# Genomic surveillance of SARS-CoV-2 in a university community: insights into tracking variants, transmission, and spread of Gamma (P.1) variant

**DOI:** 10.1101/2022.02.25.22271521

**Authors:** Ilinca I. Ciubotariu, Jack Dorman, Nicole M. Perry, Lev Gorenstein, Jobin J. Kattoor, Abebe A. Fola, Amy Zine, G. Kenitra Hendrix, Rebecca P. Wilkes, Andrew Kitchen, Giovanna Carpi

## Abstract

Using a combination of data from routine surveillance, genomic sequencing, and phylogeographic analysis we tracked the spread and introduction events of SARS-CoV-2 variants focusing on a large university community. Here, we sequenced and analyzed 677 high-quality SARS-CoV-2 genomes from positive RNA samples collected from Purdue University students, faculty, and staff who tested positive for the virus between January 2021 and May 2021, comprising an average of 32% of weekly cases across the time frame. Our analysis of circulating SARS-CoV-2 variants over time revealed periods when Variant of Concern (VOC) Alpha (B.1.1.7) and Iota (B.1.526) reached rapid dominance and documented that VOC Gamma (P.1) was increasing in frequency as campus surveillance was ending. Phylodynamic analysis of Gamma genomes from campus alongside a subsampling of >20,000 previously published P.1 genomes revealed ten independent introductions of this variant into the Purdue community, predominantly from elsewhere in the United States, with introductions from within the state of Indiana and from Illinois, and possibly Washington and New York, suggesting a degree of domestic spread. We conclude that a robust and sustained active and passive surveillance program coupled with genomic sequencing during a pandemic offers important insights into the dynamics of pathogen arrival and spread in a campus community and can help guide mitigation measures.

## BACKGROUND

Since the first severe acute respiratory syndrome-coronavirus 2 (SARS-CoV-2) cases were identified in Wuhan, Hubei Province, China, the pandemic has resulted in more than 404 million confirmed cases worldwide and over 5.7 million deaths, with the United States surpassing 77 million cases and 912,000 deaths as of February 10, 2022 (https://coronavirus.jhu.edu/map.html) [1-3]. The coronavirus disease 2019 (COVID-19) pandemic continues to create public health challenges and stress societies across the globe, which makes sustained research into viral transmission and the efficacy of community interventions particularly important.

One of the tools that has contributed to rapid progress in studying SARS-CoV-2 is genomic sequencing. This technology has led to the examination of SARS-CoV-2 global diversity and the identification of genome variants that may affect viral transmission, particularly regarding increased transmission efficacy in humans [4-6]. The Centers for Disease Control and Prevention (CDC) undertook the task of monitoring emerging SARS-CoV-2 variants and, together with the SARS-CoV-2 Interagency Group (SIG), established a classification scheme for new variants: 1) Variant of Interest (VOI), 2) Variant of Concern (VOC), and 3) Variant of High Consequence (VOHC) based on factors such as transmissibility, neutralization by antibodies, etc. [7]. As of 23 September 2021, the SIG has created a new class of variants designated as Variants Being Monitored (VBM), which include variants with substitutions of concern and variants that were previously designated as VOC or VOI that have decreased in prevalence in the United States [8]. Even so, reclassified VBMs like Gamma variant warrant continued surveillance as their roles in transmission have not yet been fully understood and thus may play important roles in the future emergence of SARS-CoV-2 variants of greater concern.

Early epidemiological studies identified that individuals with increased risk of developing severe illness or dying from COVID-19 include adults past the age of 65, individuals with pre-existing conditions like cancer, diabetes, and cardiovascular disease, and pregnant people [9]. As public health efforts to mitigate the effects of the SARS-CoV-2 pandemic were implemented in the public at large, significant efforts were made to protect the population of individuals at greatest risk for severe disease and death in particular. However, only a few months into the pandemic, during the summer of 2020, SARS-CoV-2 incidence was highest among individuals aged 20-29 years [10], possibly reflecting the efficacy of earlier interventions to protect high risk populations. Concerningly, though the risk of severe COVID-19 in young adults is relatively low compared to vulnerable individuals, the increase in cases amongst the 20-29-years-old cohort coincided with the seasonal return of students to college campuses. Of particular concern was the potential for colleges and universities to be sites of increased viral transmission that could contribute to superspreading events and community transmission into previously protected high-risk populations through networks of close contacts.

Large universities like Purdue University were inevitably going to experience SARS-CoV-2 cases and transmission during the pandemic, as the campus has an enormous student population of over 46,000 undergraduate, graduate, and professional students and over 2,400 instructional employees, as well as congregate living (e.g. student housing) and hundreds of clubs with social activities [11]. After the university took immediate action in suspending international travel in early March 2020 per the CDC guidelines, remote learning was rapidly implemented to mitigate viral spread amongst students and staff [12]. With additional safe health protocols in place, the Fall 2020 semester offered students the option of transitioning to in-person classes or continuing remotely until the following Spring, with 88% of students choosing to return to campus. The large demand for in-person instruction presented a challenge to institutional efforts to halt viral spread on campus.

During the summer, the university devised a comprehensive surveillance program coined ‘Protect Purdue Plan’, with the ultimate goal of monitoring the health of the campus community and limiting the spread of COVID-19 [13]. This plan included multiple components ranging from testing prior to arrival on campus, de-densification of academic and living spaces, and an ongoing passive surveillance testing program for on-campus students and employees via contact tracing and testing of symptomatic individuals. The plan also included active surveillance with weekly random testing by anterior nasal swabs and RT-PCR of approximately 10% of the on-campus student and employee population – a combination of mitigation strategies that were readily applied in many institutions with some degree of in-person instruction [14]. Importantly, however, little genomic evidence has been collected to understand campus transmission in general and the effects of mitigation efforts in particular [14-17].

While rapid implementation of SARS-CoV-2 genome sequencing has proven useful in the investigation of COVID-19 dynamics in other institutional settings like healthcare, few studies have been conducted on university campuses [15-23]. Some studies in the university setting have evaluated testing programs to understand control of SARS-CoV-2 transmission, while others used modeling approaches to infer SARS-CoV-2 transmission or estimate the introduction and growth rate of particular variants [17, 24-26]. Limited studies of this scale so far have investigated the dynamics of variants in a campus population over the course of a semester of in-person instruction. Here, we attempt to fill that gap and monitor SARS-CoV-2 variants and their introductions in the university population. We also argue that there is need for enhanced genomic surveillance to monitor virus lineage circulation at the local scale, especially in campus communities as the virus population will continue to evolve over time [27].

## METHODS

### Ethics Statement

The Institutional Review Board from the Purdue University Human Research Protection Program determined that viral genome sequencing of de-identified remnant COVID-19 samples included in this study is not research involving human subjects (IRB-2021-438).

### Specimen collection, RNA extraction, testing, and sampling strategies

In brief, individuals who were chosen for random campus surveillance, who had COVID-19-like symptoms, or who had been in contact with a positive case, presented themselves at various campus locations and testing occurred by way of anterior nasal swabs. The anterior nasal swabs were collected in PrimeStore^**®**^ MTM (molecular transport media) (Longhorn Vaccines & Diagnostics, Bethesda, MD) which safely inactivates infectious agents and stabilizes and preserves the released RNA for further downstream molecular applications [28].

Nucleic acid was extracted with the MagMAX™ Viral/Pathogen Nucleic Acid Isolation Kit (Applied Biosystems™, Thermo Fisher Scientific, Waltham, MA) with a KingFisher™ Flex Purification System (Thermo Fisher Scientific). Individual status (symptomatic vs. asymptomatic) based on individual report and other metadata, like travel history or vaccination status (when applicable), were noted at the time of sample collection. Collected samples were submitted to the Animal Disease Diagnostic Lab (ADDL) at Purdue University for testing, which performed nucleic acid RT-PCR using the Thermo Fisher TaqPath COVID-19 Combo Kit (Applied Biosystems™, Thermo Fisher Scientific) on a 7500 Fast Real-Time PCR System (Applied Biosystems™, Thermo Fisher Scientific). The ADDL is CLIA certified to perform high complexity testing.

Viral whole genome sequencing was performed in the Carpi Laboratory on a subset of TaqPath COVID-19 RT-PCR positive samples, based on the following two sampling strategies: 1) weekly samples that showed RT-PCR results with SGTF (TaqPath COVID-19 RT-PCR positive samples with N or ORF1AB Ct < 30 and S gene undetermined), and randomly selected positive samples with Ct values of 30 or less; and 2) retrospective sampling of randomly chosen positive samples that were not indicative of SGTF to achieve at least 20% of weekly cases. The overall goal was to conduct viral whole genome sequencing of at least 20% of weekly TaqPath COVID-19 RT-PCR positive samples from the active and passive campus surveillance schemes. The selected time frame was the first week of January 2021, through the first week of May 2021, for a total of 18 weeks. While earlier sampling could have potentially been done, testing was limited during Fall semester of 2020, and most tests were saliva-based rather than anterior nasal swabs. During the first 14 weeks of this time frame, we included all identified SGTF samples in addition to random samples, while the remaining four weeks we only included a subset of the identified SGTF samples. Retrospective sequencing included random sampling for the weeks during which we had not sequenced at least 20% of weekly cases.

### Oxford Nanopore library preparation and sequencing

The quality of a subset of the acquired RNA samples was assessed by examining any presence of RNA degradation using TapeStation High Sensitivity RNA ScreenTape (Agilent 4200, Santa Clara, CA). RNA extracts from positive samples served as an input for an amplicon-based approach for SARS-CoV-2 whole-genome sequencing on the Oxford Nanopore Technologies platforms (ONT; Oxford, United Kingdom). In brief, cDNA and amplicon libraries were generated using the “PCR tiling of SARS-CoV-2 virus” protocol (version: PTC_9096_v109_revL_06Feb2020) (Oxford Nanopore Technologies, ONT, UK). This protocol employs ARTIC V3 primers (IDT) for generating 98 amplicons of 400 bp each [29]. Sequencing libraries were prepared using the ONT Ligation Sequencing Kit (SQK-LSK109) and the Native Barcoding Expansion 1-12 and 13-24 kits for multiplexing samples, following the remaining protocol outlined in “PCR tiling of SARS-CoV-2 virus”. DNA yield following PCR amplification was assessed, with samples of concentration >20 ng/uL, as determined by Qubit dsDNA HS Assay Kit (Invitrogen, Carlsbad, CA), included in sequencing runs. No-template controls were introduced for each run at the cDNA synthesis and amplicon synthesis steps and were taken through the entire library preparation and sequencing protocol to detect any cross-contamination. The first libraries were sequenced using the MinION Mk1B platform after which all other libraries were loaded and sequenced on the GridIONX5 on R9.4.1 flow cells (Oxford Nanopore Technologies, ONT, UK) [38, 39].

### Bioinformatics processing

High accuracy basecalling were performed with Guppy basecaller version 3.1.5 on MinIT for the sequencing data generated on the MinION, and with Guppy version 4.2.4 for sequencing runs performed on the GridION5X. The resulting FASTQ files were input into the ARTIC Network bioinformatics pipeline version 1.1.3 (https://github.com/artic-network/artic-ncov2019) to generate consensus genomes using a customized pipeline on the Purdue High Performance Computing cluster [30]. Repeatable and reproducible Anaconda-based software environments for data processing were created with the condaenv-mod tool (https://github.com/amaji/conda-env-mod) [31]. Coverage plots and BAM files were visually screened as a quality control measure on randomly selected samples, the latter using Integrative Genomics Viewer (IGV) version 2.8.13 [32]. Customized scripts were used to calculate quality metrics, such as percentage of the genome sequenced and coverage depth, and sequence quality check was performed (using https://clades.nextstrain.org/) [33]. Viral genome consensus sequences were classified and lineages were assigned using the latest PANGOLIN version 3.1.17 [34]. Viral genome sequences were considered adequate for further analyses and data released to GISAID if they had >94% of the genome with 50X coverage.

### Selection of other data for context analyses

Data of positive cases for Tippecanoe County, Indiana where Purdue University is located were acquired from https://www.coronavirus.in.gov/indiana-covid-19-dashboard-and-map/ and summarized to place university cases in local context. Furthermore, to compare patterns of lineages from sequenced cases in the Purdue campus, we also used publicly available sequences on GISAID from Notre Dame University which is in South Bend, Indiana, from the University of Michigan (in a contiguous state), and from the overall state of Indiana.

### Phylogeographic analysis of P.1 and selection of context samples

For this analysis, all samples from January to May 2021 that were assigned through the bioinformatics pipeline as P.1 variant from the campus population were used, yielding a total of 18 samples. We then placed these genomes in the context of all publicly available P.1 variant genomes from the GISAID at the time this analysis was started (July 2021), or approximately 21,000 genomes. All downloaded genomes were aligned to the Purdue P.1 sequences using MAFFT with genomic data combined into a single multi FASTA file using custom scripts [35, 36]. The collection of P.1 genomes from GISAID were then screened to identify those closest to the 18 Purdue P.1 genomes. A total of 916 P.1 genomes from GISAID were within 0 to 2 substitutions of the most similar Purdue P.1 genome. After removing GISAID P.1 genomes that were identical by both sequence and location or belonging to strongly supported monophyletic groups identified in preliminary analyses that did not include Purdue sequences, the resulting dataset of 748 P.1 GISAID genomes from 41 U.S. states, Washington, D.C., and 2 other countries (Brazil and Colombia) was used to contextualize the Purdue genomes in subsequent phylogeographic analysis.

Bayesian analyses were conducted in BEAST v1.10.4 using discrete phylogeography, HKY+G nucleotide substitution, constant population size, and strict molecular clock models [37, 38]. Initial analyses were performed without biogeographic models to estimate substitution rate parameters, which, along with the overall substitution rate (8.1 × 10^−3^ sub./site/year), were fixed in the final phylogeographic inference [39]. Trees were visualized in FigTree v1.4.4 (https://github.com/rambaut/figtree/releases). Initial analyses of phylogeography were performed using Bayesian Tip-association Significance testing package (BaTS) [40]. The posterior distribution of trees from the initial runs were subjected to parsimony score calculations in BaTS, with each genome coded as either ‘Purdue’ or ‘not-Purdue’ to estimate the number of independent introductions to Purdue and as ‘Purdue’, ‘non-Purdue’, or ‘state X’ to estimate whether sequences from state X are associated with P.1 sequences from Purdue. These were complemented by subsequent Bayesian phylogenetic analyses performed in BEAST with a discrete phylogeographic model and Bayesian stochastic search variable selection to determine migration rates between Purdue and other geographic areas represented in our final sample of closely related P.1 genomes.

### Statistical Analysis

To understand the relationship between sample CT values and success in sequencing based on our threshold, we compared CT values for aggregated variants between failed and successful samples. We also used the Wilcoxon rank-sum test to assess the relationship between CT and infection status. We corrected *p*-values across the comparisons using the Benjamini-Hochberg procedure to decrease the false discovery rate [41]. When looking at the variation by U.S. state in the number of genomes deposited in GISAID, we calculated Pearson correlation coefficients.

### Data availability

All of the genomic data generated from our lab used in this research is available on GISAID (see table S3 for accession numbers). We also gratefully acknowledge the authors and submitting laboratories that generated and shared SARS-CoV-2 viral genomes via the GISAID Initiative, on which this research is based. De-identified input data are available upon request.

## RESULTS

### SARS-CoV-2 Molecular Testing and Sequencing

A total of 96,819 RT-PCR tests of in-person students and employees were performed by the ADDL facility between January 2021 and the first week of May 2021, with a positivity rate of 2.5% (2436 positive cases identified) (Table 1). Collection week 1 was defined as January 3-January 9, 2021, and this weekly enumeration continued until the last week of sample collection and sequencing, which was week 18 with corresponding dates of May 2-May 8, 2021. There was a median of 6,371 RT-PCR tests performed per week (range from 1809-7796 in weeks 4 and 6, respectively). As a result, the number of positive cases fluctuated per week, from a high of 234 during week 5 to a low of 30 positive cases in week 18 (the last week of campus active testing), as the semester ended and campus vaccination rates were picking up, although routine surveillance continued through the summer and fall semester for unvaccinated individuals (Table 1).

**Table 1.**
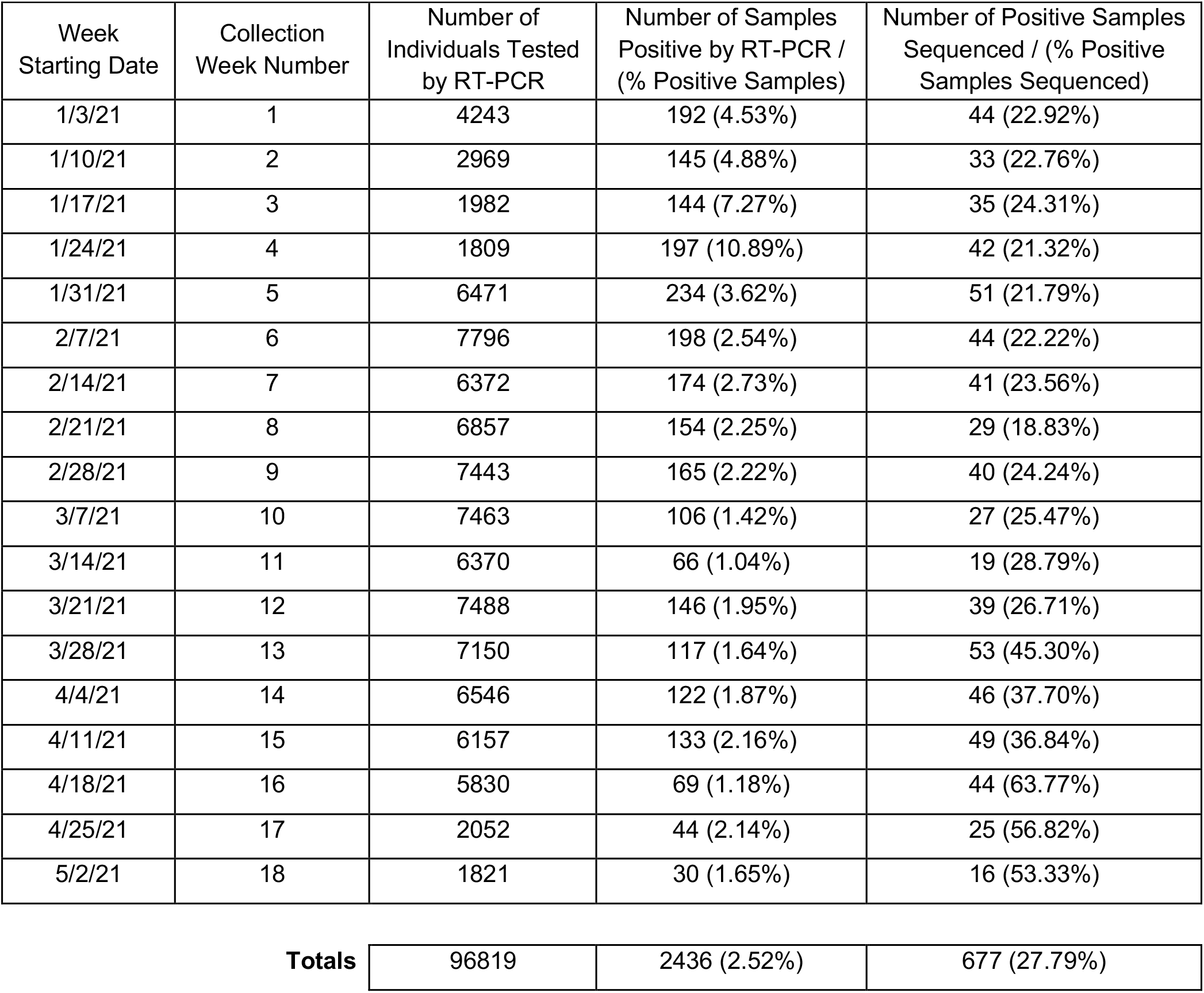
Weekly breakdown of samples tested by RT-PCR and respective results, and number of positive samples successfully sequenced from January 3, 2021-May 8, 2021, at Purdue University

The positive SARS-CoV-2 cases by RT-PCR in the campus community were placed in local context with Tippecanoe County, and it was found that Purdue accounted for roughly 35% of all positive cases during the selected time frame, with a range from a low of 15.5% in the first week to a high of 69.0% during week 9 (Fig. 1A).

**Figure 1.**
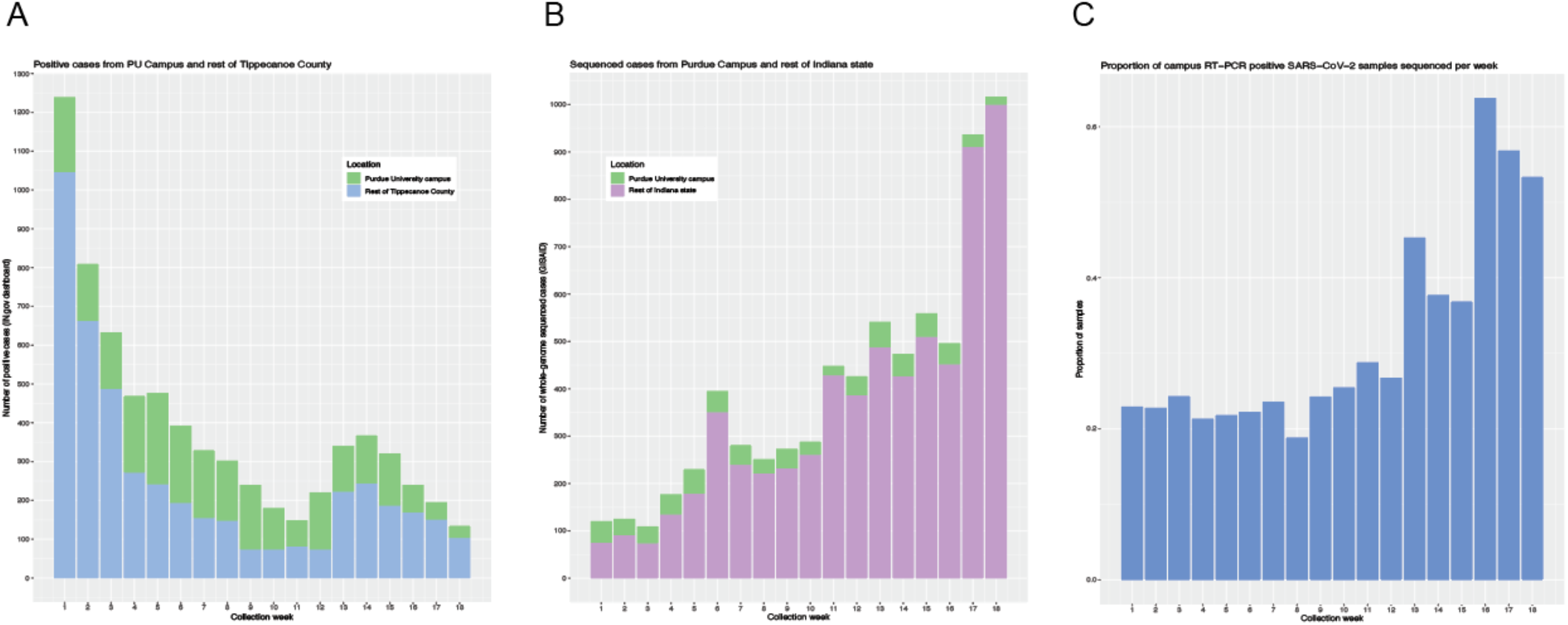
Positive cases and whole-genome sequenced samples in the laboratory in context of county positive cases, campus positive cases, and cases sequenced in the state of Indiana. *(A)* Distribution of positive SARS-CoV-2 cases from the laboratory placed in context of all positive cases in the same area as the university, Tippecanoe County, during the 18-week time frame. *(B)* Number of whole-genome sequenced SARS-CoV-2 cases from the laboratory placed in context of all sequenced cases in the state of Indiana during the 18-week time frame. *(C)* Proportion of campus RT-PCR positive SARS-CoV-2 samples among students and employees on campus from the first week of January through the first week of May 2021 that were successfully whole-genome sequenced.

We performed whole-genome sequencing on 735 samples, of which 677 (92.1%) generated high quality whole-genome sequences (at least 94% of the genome with 50X). Of the 677 samples, 431 samples were sequenced in real-time on a weekly basis to provide Protect Purdue with actionable information, and 246 were sequenced retrospectively to achieve the goal of at least 20% weekly cases. The successfully sequenced samples were submitted to GISAID and accounted for a total of 27.8% of all samples characterized as positive by RT-PCR tested by ADDL during the 18-week period (Table 1).

We aimed to contribute to the sequencing efforts of the state of Indiana, especially in the early weeks of this study when few laboratories were conducting weekly SARS-CoV-2 whole-genome sequencing (Fig. 1B). Specifically, in the first three weeks, we conducted about 32% of the sequencing in the state, though there were notably fewer positive SARS-CoV-2 cases during this time (Fig. 1B). As the rate of sequencing picked up in the state, we still contributed approximately 10% of the sequencing weekly, until the last two weeks when campus cases decreased (Fig. 1B). During this time frame, there are no other publicly available sequences in the GISAID database from this area, so we concluded that we performed the only sequencing of samples from Tippecanoe County in which the university is located. Accounting for all weeks included in this study, we sequenced an average of 32% positive cases per week (range: 19%-64% of positive samples per week) (Fig. 1C).

Although we successfully sequenced over 90% of the samples we attempted, we performed a closer analysis to understand potential reasons for failure. Here, we analyzed CT values of ORF1ab and N genes (S target values were not included here due to SGTF and thus some RT-PCR results from variants like Alpha presenting with values of 0) and identified two trends. First, when comparing the failed and successful samples, we found that the unsuccessful samples had higher median CT values (27.5) of the ORF1ab genes than the samples that generated high quality (17.6) genomes (p<0.05), which translates to the presence of a lower viral load (Supplementary Fig. 1A). Secondly, we analyzed the median N gene CT values and made a similar observation that the overall median was higher (p<0.05) for samples that failed (27.8) to generate what we considered high-quality sequences on a threshold designated as at least 94% of the genome with 50X coverage when compared to successful sequences (18.2) (Supplementary Fig. 1B).

### Asymptomatic vs. Symptomatic Infection Status

To further understand campus transmission of SARS-CoV-2 we compared asymptomatic vs. symptomatic patient status as it relates to the proportion of positive tests. Among the successfully sequenced samples, there were 41% (278) asymptomatic cases and 57% (389) symptomatic cases, with 10 cases that had unreported patient status. This pattern held true for most weeks of the study, with only three weeks (2, 11, and 15) having a greater percentage of asymptomatic cases than symptomatic cases (Supplementary Fig. 2A). We also compared the prevalence of asymptomatic and symptomatic cases among lineages identified by the WHO as variants of concern, interest, or variants being monitored (Supplementary Fig. 2B). For all of the identified lineages, there were more observed symptomatic cases than asymptomatic cases, although the differences in each respective variant were not significant (Supplementary Fig. 2B). We assumed an equal probability of an infected individual displaying symptoms (or not) and examined a binomial distribution with success rate of 0.5 to analyze the distribution of cases asymptomatic vs. symptomatic. We noted there was an observed deviation (p<0.05), indicating there was asymmetry in the distribution and more symptomatic cases. This is likely due to more symptomatic cases being identified through individuals presenting themselves at the testing facility and further undercounting asymptomatic cases as these individuals are not routinely screened. It is of note that here we aggregated all data from both sampling efforts of the active and passive surveillance.

**Figure 2.**
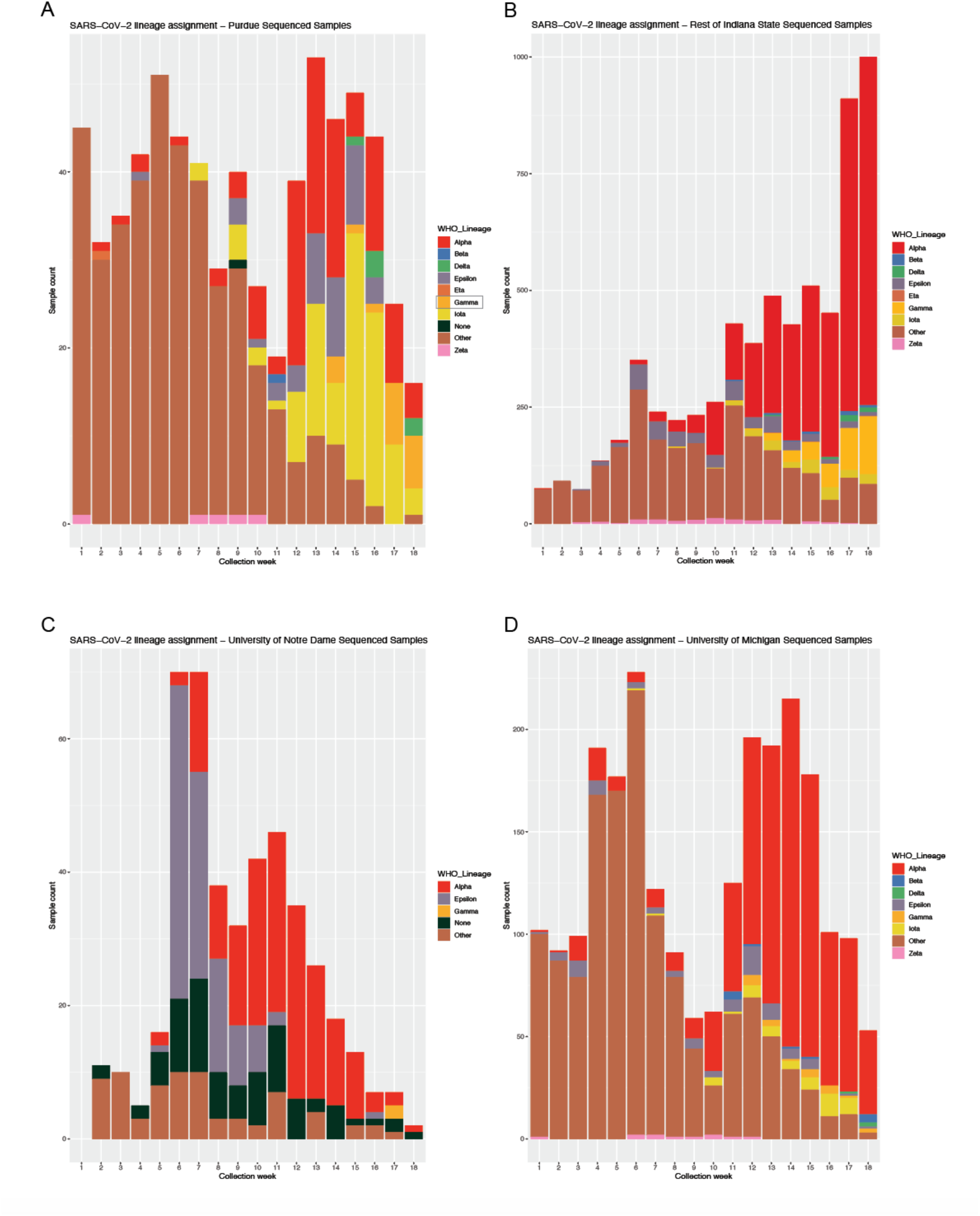
Distribution of SARS-CoV-2 WHO lineages classified as VOC and VBM (previously VOI or VOC) identified weekly from first week of January through the first week of May 2021 in four different locations. Each color and shade represent a distinct variant. Asterisk denotes week 14 when our sampling changed from a focus on SGTF cases to all random sampling. Gamma variant is highlighted in the key as subsequent phylogeographic analyses focused on this variant. *(A)* Number of cases of variants classified as VOC and VBM (previously VOI or VOC) variants per week in sequenced samples from Purdue University. Each distinct color represents one variant, while the brown color represents all “other” variants that do not fall in the category of VOC or VBM as of September 23, 2021, per the CDC. *(B)* Number of cases of variants classified as VOC and VBM in sequenced samples from the rest of Indiana state as available on GISAID from the same time period. *(C)* Number of cases of variants classified as VOC and VBM in sequenced samples from the University of Michigan as available on GISAID from the same time period. *(D)* Number of cases of variants classified as VOC and VBM in sequenced samples from the University of Notre Dame as available on GISAID from the same time period. Note that y-axes scales are different due to varying weekly testing numbers and samples identified as “None” by GISAID are likely due to lower quality in sequencing.

### Temporal Trends of Lineages in the Campus Community and in Context

Among the 677 samples successfully sequenced from campus, we observed a total of 36 lineages as identified by Pangolin, including some singleton lineages [42]. Overall, the most common lineage identified on campus was B.1.2 which accounted for 35.4% (240) of all sequenced infections, followed by B.1.1.7 at 16.1% (109). There were eight lineages (B.1.2, B.1.1.7, B.1.526, B.1.623, B.1.429, B.1.1.519, P.1, and B.1.234) which accounted for 90% (610) of the sequenced cases on campus. The remaining 28 identified lineages each presented in the population with a prevalence of less than ten cases during the study period.

To understand transmission patterns over time we assessed the prevalence of lineages each week (Fig. 5A). These results should be interpreted taking into account that B.1.1.7 was prioritized for sequencing until week 14 (as aforementioned in Methods), and then random sampling was prioritized. The most common lineage over the study period, B.1.2, accounted for over 50% of sequenced cases in the first 8 weeks of this study, after which its prevalence drastically reduced (Fig. S3). Similarly, lineages B.1.1.7 and B.1.526 accounted for the majority of cases in the last 7 weeks of the study period (Fig. S3). Some lineages like B.1.623 maintained low prevalence throughout the population over the course of the 18 weeks and did not increase to levels of high prevalence or at least 50% of weekly sequenced cases (Fig. S3). Week 4 showed the presence of 12 distinct SARS-CoV-2 lineages, which was the greatest number observed in a single week (Fig. S3).

The role of less prevalent lineages in overall virus transmission and spread are not well-understood, so we took a closer look at variants that have previously been characterized as VOCs and VOI, and some that have transitioned to VBMs (Fig. 2). The early weeks of the study presented with limited cases of VOCs or VBMs, but beginning with week 12, VOC and VBM variants accounted for most of the sequenced cases. Specifically, variants Alpha and Iota were the overwhelming majority of cases in these weeks (note: we focused on B.1.1.7 sequencing until week 14). While Fig. 2A may be interpreted as showing a decrease in Alpha variant in the last weeks of sequencing, our results are consistent with those of the rest of Indiana state, as SGTF cases (and hence, Alpha) continued to increase during this time, but we did not continue sequencing with a focus on B.1.1.7 cases. Lastly, Gamma variant started to make an appearance in the last weeks of surveillance.

A comparison of lineage prevalence was conducted to place Purdue patterns in context with the broader state of Indiana, with another university in the state of Indiana (Notre Dame), and another university in a contiguous midwestern state (University of Michigan) from publicly available sequences on GISAID (Fig 2B-2D). The overall pattern of sequenced cases was similar with that of University of Michigan, especially during weeks 12-18 as cases reached a low peak during the middle weeks of the testing period (Fig. 2D).

### Phylogenetic Analysis of Gamma Variant of Purdue Sequences and in Migration Context

Bayesian phylogenetic analysis using BEAST v1.10 was performed to place the Purdue P.1 genomes in context to circulating P.1 genomes from the wider community, specifically the United States and parts of South America (Figure 3). Initial analysis was used to estimate substitution model parameters for Bayesian phylogeographic analysis. Parsimony score-based analysis, performed on the posterior distribution of trees from the initial BEAST analyses in BaTS, was used to estimate the number of introductions to Purdue. The parsimony score (PS) is the number of character changes in a tree, and thus they are useful for identifying the number of transitions between geographic states. Coding the sequences as either non-Purdue or Purdue, the PS calculated was 10.0 (95% highest probability density = 10 – 10), suggesting 10 introductions of P.1 to the Purdue community.

**Figure 3.**
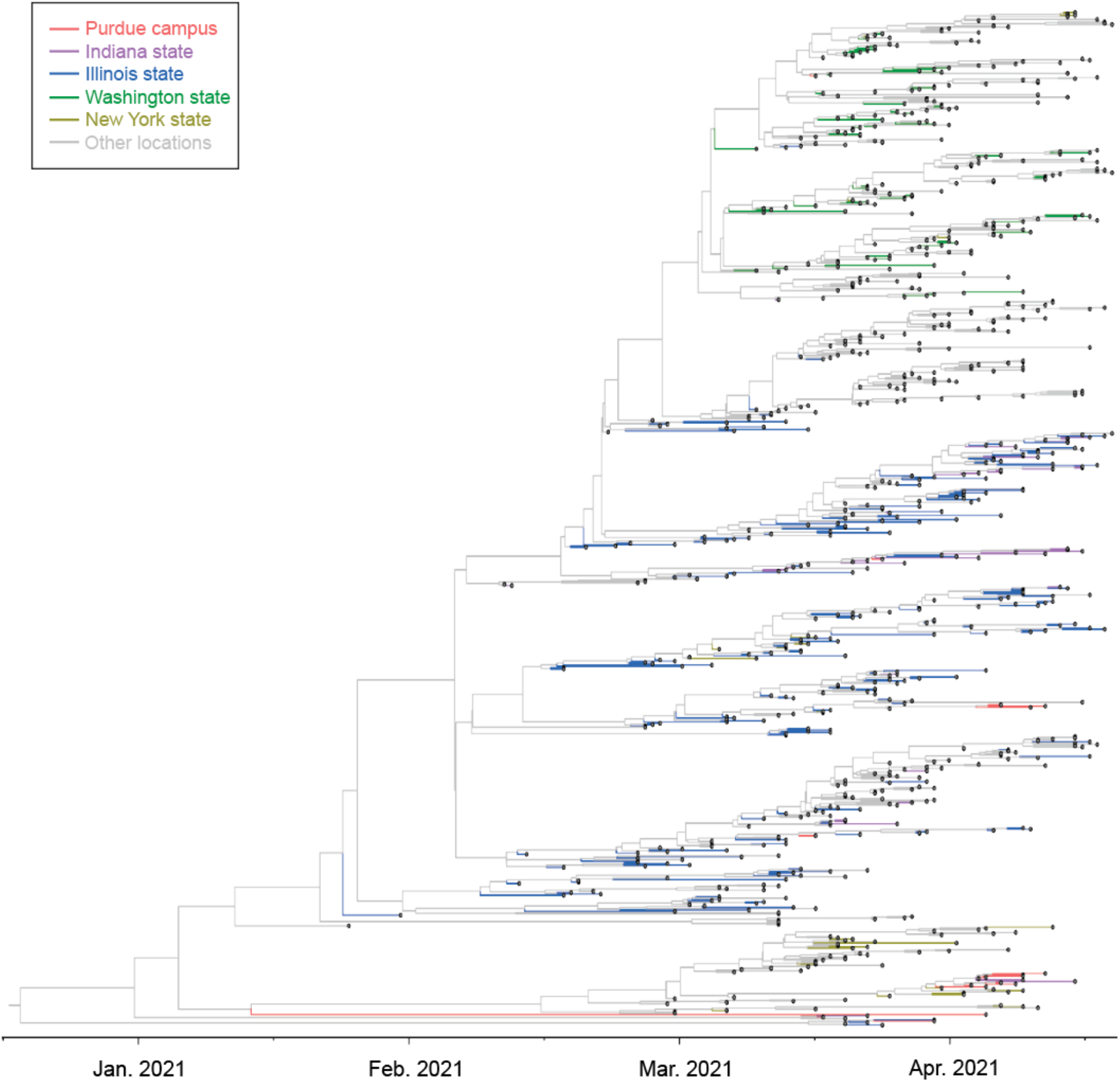
Multiple introductions, domestic spread of P.1 SARS-CoV-2 in the campus community. Time-informed Maximum Clade Credibility tree of Gamma variant (P.1) from Purdue genomes and circulating P.1 genomes from the United States and parts of South America. Included samples outside Purdue campus were downloaded from GISAID. The five colors shown (red, purple, blue, green, and yellow-green) are indicative of samples from Purdue, Indiana, Illinois, Washington, and New York respectively, with grey denoting samples from all other locations. The five colors represent the locations that had strongly supported migration rates connecting Purdue from the full model-based symmetrical discrete phylogeographic analysis.

We also performed pairwise estimates of PS scores to assess the association of Purdue sequences with sequences from other states as potential sources. The difference between parsimony scores estimated when Purdue and state X sequences are considered independently (i.e., coded separately as ‘Purdue’ and ‘state X’ categories) and when they are considered jointly (i.e., merged into one hybrid ‘Purdue-state X’ category) is an indicator of their association and evidence that state X may be a source for Purdue P.1 sequences. These values were calculated for each discrete geographic state assigned to the final sequence alignment (see Table 2). The only states with large deviations from 0 of PS values calculated independently and jointly are Illinois (4.48) and Indiana (2.78), though Washington (0.83) and New York (0.80) also had elevated PS differences; the PS differences of all other states were in the range of 0.0 – 0.25 (Supplementary Table 1).

**Table 2.** BaTS Parsimony Score

|  | Two State |  |  | Three State |
| --- | --- | --- | --- | --- |
|  | Indiana (state) | Purdue | Indiana/Purdue |  |
| Replicate 1 | 22.51 (21 – 24) | 10.00 (10 - 10) | 27.41 (26 - 29) | 30.19 (28 - 32) |
| Replicate 2 | 22.49 (21 – 24) | 10.00 (10 - 10) | 27.38 (26 - 29) | 30.17 (28 - 32) |
| Replicate 3 | 22.51 (21 – 24) | 10.00 (10 - 10) | 27.40 (26 - 29) | 30.19 (28 - 32) |
| Replicate 4 | 22.50 (21 – 24) | 10.00 (10 - 10) | 27.39 (26 - 29) | 30.18 (28 - 32) |
| Replicate 5 | 22.51 (21 – 24) | 10.00 (10 - 10) | 27.40 (26 - 29) | 30.18 (28 - 32) |

To complement the parsimony analysis of migration, we conducted full model-based symmetrical discrete phylogeographic analysis in BEAST v1.10.4. We coded all sequences as ‘Purdue’, ‘US’, ‘non-US’, or ‘state X’ to calculate migration rates between Purdue and specific geographic states in a pairwise fashion without the computationally costly overhead of estimating a full 41×41 migration matrix. Using Bayesian stochastic search variable selection and Bayes Factors (BF), we were able to identify migration rates between Purdue and particular states that had the strongest support (see Table 3) [38]. The only strongly supported migration rates connecting Purdue to individual geographic states were Purdue-Illinois (BF = 1165.01) and Purdue-Indiana (BF = 1165.01) [43]. All other migration rates between Purdue and individual states were associated with BF scores < 1.00, including Purdue-New York and Purdue-Washington, indicating no support for migration between Purdue and these regions in our dataset.

**Table 3.** BaTS Maximum Clade Size

|  | Two State |  |  | Three State |
| --- | --- | --- | --- | --- |
|  | Indiana (state) | Purdue | Indiana/Purdue |  |
| Replicate 1 | 6.09 (6 – 7) | 3.25 (3 – 5) | 11.00 (11 – 11) | Not Independent |
| Replicate 2 | 6.09 (6 – 7) | 3.24 (3 – 5) | 11.00 (11 – 11) | Not Independent |
| Replicate 3 | 6.09 (6 – 7) | 3.25 (3 – 5) | 11.00 (11 – 11) | Not Independent |
| Replicate 4 | 6.08 (6 – 7) | 3.25 (3 – 5) | 11.00 (11 – 11) | Not Independent |
| Replicate 5 | 6.09 (6 – 7) | 3.24 (3 – 5) | 11.00 (11 – 11) | Not Independent |

**Table 4.** BEAST Discrete Phylogeography 4 Rate Analysis

| State | N | Purdue-State Indicator |  | PS3 – PS2 |
| --- | --- | --- | --- | --- |
|  |  | Posterior | Bayes Factor |  |
| Alabama | 1 | 0.289 | 0.47 | 0.00 |
| Alaska | 4 | 0.272 | 0.44 | 0.00 |
| Arizona | 3 | 0.263 | 0.42 | 0.00 |
| Arkansas | 1 | 0.287 | 0.47 | 0.00 |
| California | 82 | 0.188 | 0.27 | 0.00 |
| Colorado | 3 | 0.270 | 0.43 | 0.00 |
| Connecticut | 3 | 0.270 | 0.43 | 0.00 |
| Delaware | 1 | 0.304 | 0.51 | 0.00 |
| Florida | 78 | 0.156 | 0.22 | 0.03 |
| Georgia | 11 | 0.255 | 0.40 | 0.02 |
| Hawaii | 3 | 0.283 | 0.46 | 0.00 |
| Idaho | 4 | 0.288 | 0.47 | 0.00 |
| **Illinois** | **212** | **0.999** | **1165.01** | **4.48** |
| **Indiana** | **36** | **0.999** | **1165.01** | **2.78** |
| Iowa | 2 | 0.285 | 0.46 | 0.00 |
| Kansas | 5 | 0.269 | 0.43 | 0.00 |
| Kentucky | 1 | 0.288 | 0.47 | 0.00 |
| Maryland | 3 | 0.279 | 0.45 | 0.02 |
| Massachusetts | 29 | 0.230 | 0.35 | 0.00 |
| Michigan | 14 | 0.214 | 0.32 | 0.00 |
| Minnesota | 23 | 0.226 | 0.34 | 0.02 |
| Nebraska | 4 | 0.276 | 0.44 | 0.07 |
| Nevada | 2 | 0.284 | 0.46 | 0.00 |
| New Hampshire | 1 | 0.282 | 0.46 | 0.00 |
| New Jersey | 7 | 0.254 | 0.40 | 0.05 |
| New Mexico | 1 | 0.291 | 0.48 | 0.00 |
| New York | 27 | 0.270 | 0.43 | 0.80 |
| North Carolina | 4 | 0.261 | 0.41 | 0.00 |
| Ohio | 23 | 0.249 | 0.39 | 0.00 |
| Oregon | 2 | 0.270 | 0.43 | 0.00 |
| Pennsylvania | 11 | 0.237 | 0.36 | 0.00 |
| Rhode Island | 4 | 0.255 | 0.40 | 0.00 |
| South Carolina | 10 | 0.262 | 0.41 | 0.00 |
| Tennessee | 7 | 0.261 | 0.41 | 0.00 |
| Texas | 27 | 0.214 | 0.32 | 0.17 |
| Vermont | 1 | 0.284 | 0.47 | 0.00 |
| Virginia | 1 | 0.286 | 0.47 | 0.00 |
| Washington | 68 | 0.325 | 0.56 | 0.83 |
| Wisconsin | 25 | 0.209 | 0.31 | 0.18 |
| Washington, DC | 2 | 0.277 | 0.45 | 0.00 |

## DISCUSSION

Genomic sequencing has been vital for SARS-CoV-2 surveillance and monitoring of the virus spread and evolution [44]. Our university-based SARS-CoV-2 genomic study helps inform a better understanding of community transmission at a public university with a huge population amid a pandemic. This work provides an important baseline of a SARS-CoV-2 community surveillance study following the first U.S. COVID-19 outbreak and before mass vaccination was implemented. Overall, we were able to successfully achieve viral genomic sequencing of an average of 32% of weekly cases confirmed SARS-CoV-2 positive by RT-PCR throughout the study period. This percentage is consistent with other studies conducted in a university setting, and higher than other studies done in a similarly localized population or state-level sequencing efforts in states like New York which aims to sequence 15% of weekly samples to pick up trends over time [16, 19, 20, 45]. A total of 677 whole-genomes were successfully sequenced from the Purdue campus over the course of the 18-week study period, taking into account differences in actual case number positivity and testing per week in the community.

We compared our observed WHO-denoted lineage patterns with those of other communities during the same 18 weeks based on publicly available sequences on GISAID: firstly, to the state of Indiana; secondly, to the University of Notre Dame in South Bend, Indiana; and thirdly, to the University of Michigan in Ann Arbor Michigan. We selected these three scenarios to place the Purdue lineage patterns in a broader picture, and to draw comparisons with two other universities that have performed some campus sequencing. This study sequenced 677 samples from the university from the first week of January through the first week of May 2021, the University of Notre Dame provided 448 samples, the University of Michigan had 2381 sequences, and the rest of the state of Indiana had 6,467. The number of sequenced cases for Purdue University, the University of Notre Dame, and the University of Michigan decreased greatly in the last two weeks as the semester was ending in all three locations, while the state of Indiana continued to increase the number of sequenced weekly cases. While we cannot directly compare due to lack of details regarding specific sampling strategies used in the sequencing of the samples found in GISAID, we can observe patterns based on the publicly available data. For instance, the overall picture of lineage patterns is similar among all four settings in that during the early weeks of the study period, the most dominant variants were non-VOC or VBM as characterized by the CDC. The University of Notre Dame saw the earliest shift from this pattern, with the sequenced samples showing a high percentage of VBM (namely Epsilon) in week 6. In contrast, both the state of Indiana and the University of Michigan started showing the majority presence of VBM Alpha in week 10, while Purdue University followed suit in week 12. While the overall picture of Purdue University’s lineage distribution was like that of the University of Michigan, much of the latter’s distribution was dominated by Alpha variant and other non-VBM or VOC variants, while Purdue had a great proportion of multiple VBM variants such as Iota, especially in weeks 12-18. Purdue University saw the earliest case of Alpha variant due to increased surveillance and a focus on sequencing SGTF samples, followed by the University of Michigan which saw the first sequenced case during the third week of the study period. It is possible, however, that these observed patterns are driven by individual study group sampling schemes (for instance if one cohort focused only on sequencing specific samples rather than random selection).

Variant Gamma (P.1), first documented in Brazil in November 2020, was recognized as a VOC by the US CDC and WHO and has been associated with increased transmissibility and reinfection [46-48]. In this study, it was observed that Gamma started to appear on campus in greater prevalence toward the last weeks of the surveillance period, and we sought to understand source and introduction events. Parsimony score-based phylogenetic analysis indicated that there were ten independent introductions of Gamma variant into the Purdue community during this period. Further analysis looking at other states as potential sources for Purdue Gamma variant sequences showed that the sole supported sources of introductions to the Purdue community were Illinois, Indiana, Washington, and New York. These results were also supported by a complimentary discrete phylogeographic analysis, with the highest levels of support in migration rates for Illinois and Indiana, with lower levels of support for New York and Washington. Moreover, these results are consistent with epidemiological link, with three of these cases having confirmed travel history in Illinois. However, due to the limited travel history provided by the tested individuals, we are unable to correlate all the multiple introductions as determined by the phylogeographic analysis with epidemiological and travel data. Domestic spread and multiple introductions of Gamma variant are also consistent with results from another study investigating transmission of P.1 in New England [49]. Overall, our finding reflects the fact that Purdue University has a high rate of in-state and midwestern students who likely traveled home immediately prior to or during the sampling period.

This study had some limitations, including the fact that we used two sampling schemes, as one goal was to ensure we were tracking active cases of SGTF for proper case and contact isolation, and the second was to sequence more cases to gain a better understanding of transmission. Moreover, it was not possible to compare our results to other sequencing data within the county or neighboring counties as no data were available on GISAID. Due to inadequacies of SARS-CoV-2 genomic surveillance in many places across the US, our estimates of introductions of Gamma variant to the campus may be biased toward states that had higher sequencing effort and enhanced genomic surveillance.

Indeed, there is substantial variation by U.S. state in the number of genomes deposited in the GISAID database (total n=714,368 through June 2021 with assigned U.S. state origin) (Supplementary Table 2). Through June 2021, the median number of total genome sequences deposited for U.S. states was 6910.5, with a mean of 14,287 and with a range of 732 (South Dakota) to 102988 (California). The collection of GISAID P.1/Gamma genomes available as of July 2021 was also skewed by U.S. state, with a median of 60 P.1 sequences, (range of 3 (South Dakota) to 2035 (Illinois)). However, the correlation between the total number of SARS-CoV-2 sequences with state of origins data through June 2021 in GISAID and state population sizes (U.S. Census Bureau, https://data.census.gov) is substantial (Pearson’s r = 0.91); this may indicate that variation in sequenced genomes per state is the direct product of proportional differences in state population or that proportional differences in state population size drove differences in sample collection and sequencing. The correlation with state population also holds for the total number of P.1 genomes in GISAID through June 2021 (Pearson’s r = 0.58). Interestingly, the number of samples from each state in our contextual P.1 genome dataset is also highly correlated with the number of samples from each state in the dataset of all P.1 genomes (Pearson’s r = 0.90); this is so despite the exclusion of all genomes not closely related to the sampled genomes from Purdue, which constituted only ∼4% of all P.1 sequences in GISAID. This may reflect the rapid spread of P.1 through the United States by the summer of 2021, during which the P.1 lineage accumulated diversity at a pace (0.0008 sub./site/year or ∼2 sub./month) relative to its geographic diffusion that precluded the establishment of sub-lineages associated with particularly narrow geographic distributions. Despite this, we were able to identify clear geographic patterns of relatedness between viral genomes indicative of geographic sources for P.1 genomes sampled from the Purdue community.

## CONCLUSIONS

By implementing SARS-CoV-2 genomic sequencing nested within passive and active surveillance over the course of one semester at Purdue University, we were able to investigate SARS-CoV-2 transmission dynamics in a university setting. We identified variants of differing levels of concern in the 677 newly sequenced viral genomes and compared variant temporal trends to other similar university settings and in a broader context using publicly available data. Further phylodynamic analysis of Gamma (P.1) genomes from campus revealed multiple introductions into the Purdue community, predominantly from states within the United States. We show that robust surveillance programs coupled with viral genomic sequencing and phylogenetic analysis can provide critical insights into SARS-CoV-2 transmission dynamics, variant arrival and spread in universities, and can help inform mitigation strategies for future waves, especially as SARS-CoV-2 continues to circulate and evolve.

## Data Availability

All of the genomic data generated from our lab used in this research is available on GISAID (see table S3 for accession numbers). We also gratefully acknowledge the authors and submitting laboratories that generated and shared SARS-CoV-2 viral genomes via the GISAID Initiative, on which this research is based.

## AUTHOR CONTRIBUTIONS

I.C., J.D., and G.C. designed the research study. G.C., G.K.H., and R.P.W. coordinated the sample selection. G.K.H. and R.P.W. provided samples and epidemiological data. J.D., I.C., N.P., and J.K. generated sequencing data. G.C., J.D., and L.G. performed bioinformatics analysis. A.K., I.C., A.Z. and G.C. analyzed sequencing data and performed the phylogenetic analyses. I.C. drafted the initial manuscript. All authors read, reviewed, and approved the final manuscript.

## ACKNOWLEDGEMENTS

We would like to thank Purdue University for conducting on-campus SARS-CoV-2 testing and surveillance. We also thank the ADDL staff for performing RT-PCR on all samples. Lastly, we thank the groups that continuously make their sequencing data available to the public.

## REFERENCES

1. Dong E, Du H, Gardner L. An interactive web-based dashboard to track COVID-19 in real time. Lancet Infect Dis 2020; 20:533–4.

2. Wu F, Zhao S, Yu B, et al. A new coronavirus associated with human respiratory disease in China. Nature 2020; 579:265–9.

3. Zhu N, Zhang D, Wang W, et al. A Novel Coronavirus from Patients with Pneumonia in China, 2019. N Engl J Med 2020; 382:727–33.

4. Morais IJ, Polveiro RC, Souza GM, Bortolin DI, Sassaki FT, Lima ATM. The global population of SARS-CoV-2 is composed of six major subtypes. Sci Rep 2020; 10:18289.

5. Brufsky A. Distinct viral clades of SARS-CoV-2: Implications for modeling of viral spread. J Med Virol 2020; 92:1386–90.

6. Wang C, Liu Z, Chen Z, et al. The establishment of reference sequence for SARS-CoV-2 and variation analysis. J Med Virol 2020; 92:667–74.

7. CDC. SARS-CoV-2 Variants. Available at: https://www.cdc.gov/coronavirus/2019-ncov/cases-updates/variant-surveillance/variant-info.html.

8. CDC. SARS-CoV-2 Variant Classifications and Definitions. Available at: https://www.cdc.gov/coronavirus/2019-ncov/variants/variant-info.html.

9. CDC. COVID-19 Information for Specific Groups of People. Available at: https://www.cdc.gov/coronavirus/2019-ncov/need-extra-precautions/index.html.

10. Boehmer TK, DeVies J, Caruso E, etal. Changing Age Distribution of the COVID-19 Pandemic — United States, May–August 2020. MMWR Morb Mortal Wkly Rep 2020 2020; 69:1404–9.

11. Oates M. Purdue announces fall enrollment numbers. Available at: https://www.purdue.edu/newsroom/releases/2020/Q3/purdue-announces-fall-enrollment-numbers.html.

12. Purdue. Protect Purdue COVID-19 Dashboard. Available at: https://protect.purdue.edu/timeline/.

13. Purdue. Protect Purdue Plan. Available at: https://protect.purdue.edu/plan/.

14. Walke HT, Honein MA, Redfield RR. Preventing and Responding to COVID-19 on College Campuses. JAMA 2020; 324:1727–8.

15. Hamer DH, White LF, Jenkins HE, et al. Assessment of a COVID-19 Control Plan on an Urban University Campus During a Second Wave of the Pandemic. JAMA Netw Open 2021; 4:e2116425–e.

16. Avendano C, Lilienfeld A, Rulli L, et al. SARS-CoV-2 Variant Tracking and Mitigation During In-Person Learning at a Midwestern University in the 2020-2021 School Year. JAMA Netw Open 2022; 5:e2146805–e.

17. Petros BA, Turcinovic J, Welch NL, et al. Early introduction and rise of the Omicron SARS-CoV-2 variant in highly vaccinated university populations. medRxiv 2022:2022.01.27.22269787.

18. Meredith LW, Hamilton WL, Warne B, et al. Rapid implementation of SARS-CoV-2 sequencing to investigate cases of health-care associated COVID-19: a prospective genomic surveillance study. Lancet Infect Dis 2020; 20:1263–71.

19. Valesano AL, Fitzsimmons WJ, Blair CN, et al. SARS-CoV-2 Genomic Surveillance Reveals Little Spread From a Large University Campus to the Surrounding Community. Open Forum Infect Dis 2021; 8:ofab518–ofab.

20. Aggarwal D, Fieldman T, Warne B, COG-UK, Screening UoCAC-, Consortium P. Genomic epidemiology of SARS-CoV-2 in the University of Cambridge identifies dynamics of transmission: an interim report, 10 December 2020. Scientific Advisory Group for Emergencies 2020.

21. Ramirez E, Wilkes RP, Carpi G, Dorman J, Bowen C, Smith L. SARS-Cov-2 Breakthrough Infections in Fully Vaccinated Individuals in a University Setting. Journal of Infectious Diseases & Therapy 2021; 9.

22. Rebmann T, Loux TM, Arnold LD, Charney R, Horton D, Gomel A. SARS-CoV-2 Transmission to Masked and Unmasked Close Contacts of University Students with COVID-19 - St. Louis, Missouri, January-May 2021. MMWR Morb Mortal Wkly Rep 2021; 70:1245–8.

23. Nixon E, Thomas A, Stocks D, et al. Impacts of vaccination and asymptomatic testing on SARS-CoV-2 transmission dynamics in a university setting. medRxiv 2021:2021.11.22.21266565.

24. Weil AA, Sohlberg SL, O’Hanlon JA, et al. SARS-CoV-2 Epidemiology on a Public University Campus in Washington State. Open Forum Infect Dis 2021.

25. Johnson KE, Woody S, Lachmann M, et al. Early estimates of SARS-CoV-2 B.1.1.7 variant emergence in a university setting. medRxiv 2021:2021.03.05.21252541.

26. Hill EM, Atkins BD, Keeling MJ, Tildesley MJ, Dyson L. Modelling SARS-CoV-2 transmission in a UK university setting. Epidemics 2021; 36:100476.

27. CDC. Emerging SARS-CoV-2 Variants. Available at: https://www.cdc.gov/coronavirus/2019-ncov/more/science-and-research/scientific-brief-emerging-variants.html.

28. CDC. Longhorn PrimeStore® Molecular Transport Medium Fact Sheet. Available at: https://www.cdc.gov/coronavirus/2019-ncov/downloads/lab/Longhorn-Molecular-Transport-Medium.pdf.

29. Quick J. nCoV-2019 sequencing protocol v3 (LoCost) V.3. protocolsio 2020.

30. Hacker T, Yang B, McCartney G. Empowering Faculty: A Campus Cyberinfrastructure Strategy for Research Communities. Educause Review 2014.

31. Maji AK, Gorenstein L, Lentner G. Demystifying Python Package Installation with conda-env-mod. In: 2020 IEEE/ACM International Workshop on HPC User Support Tools (HUST) and Workshop on Programming and Performance Visualization Tools (ProTools).27-37.

32. Robinson JT, Thorvaldsdóttir H, Winckler W, et al. Integrative genomics viewer. Nat Biotechnol 2011; 29:24–6.

33. Aksamentov I, Roemer C, Hodcroft EB, Neher RA. Nextclade: clade assignment, mutation calling and quality control for viral genomes. J Open Source Softw 2021; 6:3773.

34. Rambaut A, Holmes EC, O’Toole Á, et al. A dynamic nomenclature proposal for SARS-CoV-2 lineages to assist genomic epidemiology. Nat Microbiol 2020; 5:1403–7.

35. Katoh K, Standley DM. MAFFT Multiple Sequence Alignment Software Version 7: Improvements in Performance and Usability. Mol Biol Evol 2013; 30:772–80.

36. Katoh K, Misawa K, Kuma Ki, Miyata T. MAFFT: a novel method for rapid multiple sequence alignment based on fast Fourier transform. Nucleic Acids Res 2002; 30:3059–66.

37. Suchard MA, Lemey P, Baele G, Ayres DL, Drummond AJ, Rambaut A. Bayesian phylogenetic and phylodynamic data integration using BEAST 1.10. Virus Evol 2018; 4.

38. Lemey P, Rambaut A, Drummond AJ, Suchard MA. Bayesian Phylogeography Finds Its Roots. PLoS Comput Biol 2009; 5:e1000520.

39. Ghafari M, du Plessis L, Pybus O, Katzourakis A. Time dependence of SARS-CoV-2 substitution rates. https://virological.org/t/time-dependence-of-sars-cov-2-substitution-rates/542, 2020.

40. Parker J, Rambaut A, Pybus OG. Correlating viral phenotypes with phylogeny: Accounting for phylogenetic uncertainty. Infect Genet Evol 2008; 8:239–46.

41. Benjamini Y, Hochberg Y. Controlling the False Discovery Rate: A Practical and Powerful Approach to Multiple Testing. J R Stat Soc Series B Stat Methodol 1995; 57:289–300.

42. O’Toole Á, Scher E, Underwood A, et al. Assignment of epidemiological lineages in an emerging pandemic using the pangolin tool. Virus Evol 2021; 7.

43. Kass RE, Raftery AE. Bayes Factors. J Am Stat Assoc 1995; 90:773–95.

44. Morris CP, Luo CH, Amadi A, et al. An Update on Severe Acute Respiratory Syndrome Coronavirus 2 Diversity in the US National Capital Region: Evolution of Novel and Variants of Concern. Clin Infect Dis 2021.

45. Constantino AK, Kopecki D, Kimball S. New York officials confirm 5 cases of Covid omicron variant in NYC metro area. CNBC, 2021.

46. Imai M, Halfmann PJ, Yamayoshi S, et al. Characterization of a new SARS-CoV-2 variant that emerged in Brazil. Proc Natl Acad Sci 2021; 118:e2106535118.

47. Faria NR, Mellan TA, Whittaker C, et al. Genomics and epidemiology of a novel SARS-CoV-2 lineage in Manaus, Brazil. medRxiv : the preprint server for health sciences 2021:2021.02.26.21252554.

48. Fujino T, Nomoto H, Kutsuna S, et al. Novel SARS-CoV-2 Variant in Travelers from Brazil to Japan. Emerg Infect Dis 2021; 27.

49. Brown C, Doucette M, Fink T, et al. Detection of a large cluster and multiple introductions of the P.1 SARS-CoV-2 Variant of Concern in Massachusetts. https://virological.org/t/detection-of-a-large-cluster-and-multiple-introductions-of-the-p-1-sars-cov-2-variant-of-concern-in-massachusetts/671, 2021.

