## Supplementary Tables and Figures for "Genomic surveillance of SARS-CoV-2 in a university community: insights into tracking variants, transmission, and spread of Gamma (P.1) variant"

**Supplementary Table 1.** Discrete geographic state analysis with PS differences for all states

| State | N | Two State | | Three State | | PS3 – PS2 |
|---|---|---|---|---|---|---|
| | | PS2 | MC (Purdue+State) | PS3 | MC (State) | |
| Alabama | 1 | 11.00 (11 – 11) | 3.25 (3 – 5) | 11.00 (11 – 11) | 1.00 (1 – 1) | 0.00 |
| Alaska | 4 | 13.16 (12 – 14) | 3.25 (3 – 5) | 13.16 (12 – 14) | 1.71 (1 – 2) | 0.00 |
| Arizona | 3 | 13.00 (13 – 13) | 3.25 (3 – 5) | 13.00 (13 – 13) | 1.00 (1 – 1) | 0.00 |
| Arkansas | 1 | 11.00 (11 – 11) | 3.25 (3 – 5) | 11.00 (11 – 11) | 1.00 (1 – 1) | 0.00 |
| California | 82 | 60.11 (56 – 64) | 6.05 (3 – 11) | 60.11 (56 – 64) | 6.01 (3 – 11) | 0.00 |
| Colorado | 3 | 12.11 (12 – 13) | 3.25 (3 – 5) | 12.11 (12 – 13) | 1.89 (1 – 2) | 0.00 |
| Connecticut | 3 | 12.92 (12 – 13) | 3.25 (3 – 5) | 12.92 (12 – 13) | 1.08 (1 – 2) | 0.00 |
| Delaware | 1 | 11.00 (11 – 11) | 3.25 (3 – 5) | 11.00 (11 – 11) | 1.00 (1 – 1) | 0.00 |
| Florida | 78 | 69.77 (66 – 73) | 7.57 (4 – 9) | 69.80 (66 – 73) | 7.55 (4 – 9) | 0.03 |
| Georgia | 11 | 20.84 (20 – 21) | 3.25 (3 – 5) | 20.86 (20 – 21) | 1.13 (1 – 2) | 0.02 |
| Hawaii | 3 | 12.93 (12 – 13) | 3.25 (3 – 5) | 12.93 (12 – 13) | 1.06 (1 – 2) | 0.00 |
| Idaho | 4 | 13.82 (13 – 14) | 3.25 (3 – 5) | 13.82 (13 – 14) | 1.17 (1 – 2) | 0.00 |
| Illinois | 212 | 94.70 (89 – 100) | 13.80 (12 – 17) | 99.18 (94 – 104) | 13.80 (12 – 17) | 4.48 |
| Indiana | 36 | 27.41 (26 – 29) | 11.00 (11 – 11) | 30.19 (28 – 32) | 6.09 (6 – 7) | 2.78 |
| Iowa | 2 | 11.98 (12 – 12) | 3.25 (3 – 5) | 11.98 (12 – 12) | 1.018 (1 – 1) | 0.00 |
| Kansas | 5 | 14.86 (14 – 15) | 3.25 (3 – 5) | 14.86 (14 – 15) | 1.13 (1 – 2) | 0.00 |
| Kentucky | 1 | 11.00 (11 – 11) | 3.25 (3 – 5) | 11.00 (11 – 11) | 1.00 (1 – 1) | 0.00 |
| Maryland | 3 | 12.98 (13 – 13) | 3.25 (3 – 5) | 13.00 (13 – 13) | 1.00 (1 – 1) | 0.02 |
| Massachusetts | 29 | 33.53 (31 – 36) | 3.64 (3 – 5) | 33.53 (31 – 36) | 3.22 (2 – 5) | 0.00 |
| Michigan | 14 | 21.75 (21 – 22) | 3.41 (3 – 5) | 21.77 (21 – 22) | 3.17 (3 – 4) | 0.00 |

| | | | | | | |
|---|---|---|---|---|---|---|
| Minnesota | 23 | 32.06 (30 – 33) | 3.26 (3 – 5) | 32.08 (30 – 33) | 1.61 (1 – 2) | 0.02 |
| Nebraska | 4 | 13.90 (13 – 14) | 3.30 (3 – 5) | 13.97 (14 – 14) | 1.03 (1 – 1) | 0.07 |
| Nevada | 2 | 11.98 (12 – 12) | 3.25 (3 – 5) | 11.98 (12 – 12) | 1.02 (1 – 1) | 0.00 |
| New Hampshire | 1 | 11.00 (11 – 11) | 3.25 (3 – 5) | 11.00 (11 – 11) | 1.00 (1 – 1) | 0.00 |
| New Jersey | 7 | 16.89 (16 – 17) | 3.25 (3 – 5) | 16.94 (16 – 17) | 1.06 (1 – 2) | 0.05 |
| New Mexico | 1 | 11.00 (11 – 11) | 3.25 (3 – 5) | 11.00 (11 – 11) | 1.00 (1 – 1) | 0.00 |
| New York | 27 | 30.80 (28 – 33) | 3.39 (3 – 5) | 31.60 (29 – 34) | 2.63 (2 – 4) | 0.80 |
| North Carolina | 4 | 13.97 (14 – 14) | 3.25 (3 – 5) | 13.97 (14 – 14) | 1.02 (1 – 1) | 0.00 |
| Ohio | 23 | 28.84 (26 – 31) | 3.34 (3 – 5) | 28.84 (26 – 31) | 2.43 (2 – 4) | 0.00 |
| Oregon | 2 | 12.00 (12 – 12) | 3.25 (3 – 5) | 12.00 (12 – 12) | 1.00 (1 – 1) | 0.00 |
| Pennsylvania | 11 | 19.94 (19 – 20) | 3.25 (3 – 5) | 19.94 (19 – 20) | 2.01 (2 – 2) | 0.00 |
| Rhode Island | 4 | 14.00 (14 – 14) | 3.25 (3 – 5) | 14.00 (14 – 14) | 1.00 (1 – 1) | 0.00 |
| South Carolina | 10 | 19.46 (18 – 20) | 3.25 (3 – 5) | 19.46 (18 – 20) | 1.45 (1 – 1) | 0.00 |
| Tennessee | 7 | 13.98 (14 – 14) | 4.09 (4 – 5) | 13.98 (14 – 14) | 4.01 (4 – 4) | 0.00 |
| Texas | 27 | 31.67 (29 – 34) | 3.62 (3 – 5) | 31.84 (29 – 34) | 2.82 (2 – 5) | 0.17 |
| Vermont | 1 | 11.00 (11 – 11) | 3.25 (3 – 5) | 11.00 (11 – 11) | 1.00 (1 – 1) | 0.00 |
| Virginia | 1 | 11.00 (11 – 11) | 3.25 (3 – 5) | 11.00 (11 – 11) | 1.00 (1 – 1) | 0.00 |
| Washington | 68 | 63.81 (59 – 69) | 3.57 (3 – 5) | 64.66 (60 – 69) | 3.11 (2 – 5) | 0.83 |
| Wisconsin | 25 | 30.54 (29 – 31) | 5.00 (5 – 5) | 30.72 (30 – 31) | 4.99 (5 – 5) | 0.18 |
| Washington, DC | 2 | 12.00 (12 – 12) | 3.25 (3 – 5) | 12.00 (12 – 12) | 1.00 (1 – 1) | 0.00 |
| Brazil | 1 | 11.00 (11 – 11) | 3.25 (3 – 5) | 11.00 (11 – 11) | 1.00 (1 – 1) | 0.00 |
| Colombia | 1 | 10.75 (10 – 11) | 3.25 (3 – 5) | 11.00 (11 – 11) | 1.00 (1 – 1) | 0.25 |

**Supplementary Table 2**. Dataset sample sizes, population sizes, and Bayes Factor support for migration by state

| | Population[1] | Total Genomes[2] | Total P.1 Genomes[3] | Contextual P.1 Genomes[4] | Subsampled Contextual P.1 Genomes[5] | Migration Bayes Factor[6] |
|---|---|---|---|---|---|---|
| Alabama | 5039877 | 3640 | 29 | 2 | 1 | 0.47 |
| Alaska | 732673 | 2636 | 16 | 4 | 4 | 0.44 |
| Arizona | 7276316 | 23711 | 203 | 7 | 3 | 0.42 |
| Arkansas | 3025891 | 1985 | 16 | 1 | 1 | 0.47 |
| California | 39237836 | 102988 | 1119 | 86 | 82 | 0.27 |
| Colorado | 5812069 | 30037 | 184 | 13 | 3 | 0.43 |
| Connecticut | 3605597 | 8485 | 107 | 3 | 3 | 0.43 |
| Delaware | 1003384 | 2536 | 29 | 1 | 1 | 0.51 |
| Florida | 21781128 | 45773 | 1472 | 99 | 78 | 0.22 |
| Georgia | 10799566 | 8691 | 89 | 11 | 11 | 0.40 |
| Hawaii | 1441553 | 2989 | 38 | 3 | 3 | 0.46 |
| Idaho | 1900923 | 2748 | 10 | 4 | 4 | 0.47 |
| Illinois | 12671469 | 21708 | 2035 | 262 | 212 | 1165.01 |
| Indiana | 6805985 | 8931 | 336 | 75 | 54 | 1165.01 |
| Iowa | 3193079 | 1115 | 22 | 2 | 2 | 0.46 |
| Kansas | 2934582 | 4271 | 129 | 7 | 5 | 0.43 |
| Kentucky | 4509394 | 3139 | 27 | 1 | 1 | 0.47 |
| Louisiana | 4624047 | 6373 | 20 | 0 | 0 | NA |
| Maine | 1372247 | 5141 | 53 | 0 | 0 | NA |
| Maryland | 6165129 | 15675 | 57 | 3 | 3 | 0.45 |
| Massachusetts | 6984723 | 21638 | 1180 | 35 | 29 | 0.35 |
| Michigan | 10050811 | 29428 | 258 | 14 | 14 | 0.32 |

| | | | | | | |
|---|---|---|---|---|---|---|
| Minnesota | 5707390 | 27600 | 278 | 30 | 23 | 0.34 |
| Mississippi | 2949965 | 1326 | 5 | 0 | 0 | NA |
| Missouri | 6168187 | 5092 | 69 | 0 | 0 | NA |
| Montana | 1104271 | 2223 | 8 | 0 | 0 | NA |
| Nebraska | 1963692 | 3218 | 21 | 5 | 4 | 0.44 |
| Nevada | 3143991 | 6334 | 26 | 2 | 2 | 0.46 |
| New Hampshire | 1388992 | 2350 | 62 | 1 | 1 | 0.46 |
| New Jersey | 9267130 | 16211 | 230 | 12 | 7 | 0.40 |
| New Mexico | 2115877 | 7190 | 30 | 3 | 1 | 0.48 |
| New York | 19835913 | 51873 | 438 | 31 | 27 | 0.43 |
| North Carolina | 10551162 | 11885 | 82 | 4 | 4 | 0.41 |
| North Dakota | 774948 | 3350 | 9 | 0 | 0 | NA |
| Ohio | 11780017 | 11168 | 190 | 26 | 23 | 0.39 |
| Oklahoma | 3986639 | 915 | 5 | 0 | 0 | NA |
| Oregon | 4246155 | 12170 | 185 | 3 | 2 | 0.43 |
| Pennsylvania | 12964056 | 18950 | 171 | 11 | 11 | 0.36 |
| Rhode Island | 1095610 | 4352 | 148 | 7 | 4 | 0.40 |
| South Carolina | 5190705 | 4159 | 46 | 10 | 10 | 0.41 |
| South Dakota | 895376 | 732 | 3 | 0 | 0 | NA |
| Tennessee | 6975218 | 5203 | 62 | 7 | 7 | 0.41 |
| Texas | 29527941 | 63090 | 438 | 28 | 27 | 0.32 |
| Utah | 3337975 | 19777 | 16 | 0 | 0 | NA |
| Vermont | 645570 | 1246 | 10 | 1 | 1 | 0.46 |
| Virginia | 8642274 | 10300 | 46 | 2 | 1 | 0.47 |
| Washington | 7738692 | 32903 | 1035 | 80 | 68 | 0.56 |
| West Virginia | 1782959 | 6631 | 11 | 0 | 0 | NA |
| Wisconsin | 5895908 | 16916 | 179 | 33 | 25 | 0.31 |
| Wyoming | 578803 | 13566 | 21 | 0 | 0 | NA |

1 = estimates of 2021 state populations from the U.S. Census Bureau, https://data.census.gov

2 = total number of SARS-CoV-2 genomes sampled through June 2021 available on GISAID from each state

3 = total number of variant P.1/Gamma genomes sampled through June 2021 available on GISAID from each state

4 = number of variant P.1/Gamma genomes within 0-2 substitutions of Purdue available on GISAID from each state

5 = number of variant P.1/Gamma genomes by state used in phylogeographic analysis after sub-sampling all available P.1 genomes within 0-2 substitutions of Purdue genomes

6 = Bayes Factors supporting migration between each state and Purdue estimated from phylogenetic analysis of GISAID genomes closely related to Purdue P.1 samples

**Supplementary Table 3**. All 677 sequenced samples collected at Purdue University campus from January 3-May 8, 2021, included in the study with respective virus names and accession IDs after submission to GISAID

| GISAID Virus Name | GISAID Accession ID |
|---|---|
| hCoV-19/USA/IN-PU-82386/2021 | EPI_ISL_893214 |
| hCoV-19/USA/IN-PU-100821/2021 | EPI_ISL_891144 |
| hCoV-19/USA/IN-PU-104025/2021 | EPI_ISL_891141 |
| hCoV-19/USA/IN-PU-LH000094162/2021 | EPI_ISL_955222 |
| hCoV-19/USA/IN-PU-LH000101146/2021 | EPI_ISL_955226 |
| hCoV-19/USA/IN-PU-LH000094074/2021 | EPI_ISL_955220 |
| hCoV-19/USA/IN-PU-LH000124690/2021 | EPI_ISL_955224 |
| hCoV-19/USA/IN-PU-LH000145185/2021 | EPI_ISL_955215 |
| hCoV-19/USA/IN-PU-LH000115045/2021 | EPI_ISL_955219 |
| hCoV-19/USA/IN-PU-LH000094242/2021 | EPI_ISL_955221 |
| hCoV-19/USA/IN-PU-LH000124570/2021 | EPI_ISL_955223 |
| hCoV-19/USA/IN-PU-LH000092865/2021 | EPI_ISL_955228 |
| hCoV-19/USA/IN-PU-LH000046230/2021 | EPI_ISL_955225 |
| hCoV-19/USA/IN-PU-LH000100666/2021 | EPI_ISL_955217 |
| hCoV-19/USA/IN-PU-LH000109094/2021 | EPI_ISL_955216 |
| hCoV-19/USA/IN-PU-LH000145107/2021 | EPI_ISL_955218 |
| hCoV-19/USA/IN-PU-LH000092821/2021 | EPI_ISL_955227 |
| hCoV-19/USA/IN-PU-LH000101372/2021 | EPI_ISL_955212 |
| hCoV-19/USA/IN-PU-LH000115377/2021 | EPI_ISL_1184180 |
| hCoV-19/USA/IN-PU-LH000132511/2021 | EPI_ISL_1184177 |
| hCoV-19/USA/IN-PU-LH000121973/2021 | EPI_ISL_1184183 |
| hCoV-19/USA/IN-PU-LH000145467/2021 | EPI_ISL_1184172 |

| | |
|---|---|
| hCoV-19/USA/IN-PU-LH000115449/2021 | EPI_ISL_1184169 |
| hCoV-19/USA/IN-PU-LH000094402/2021 | EPI_ISL_1184174 |
| hCoV-19/USA/IN-PU-LH000115260/2021 | EPI_ISL_1184170 |
| hCoV-19/USA/IN-PU-LH000145457/2021 | EPI_ISL_1184171 |
| hCoV-19/USA/IN-PU-LH000094397/2021 | EPI_ISL_1184175 |
| hCoV-19/USA/IN-PU-LH000121610/2021 | EPI_ISL_1184176 |
| hCoV-19/USA/IN-PU-LH000117345/2021 | EPI_ISL_1184178 |
| hCoV-19/USA/IN-PU-LH000014094/2021 | EPI_ISL_1184179 |
| hCoV-19/USA/IN-PU-LH000122006/2021 | EPI_ISL_1184184 |
| hCoV-19/USA/IN-PU-LH000145283/2021 | EPI_ISL_1184173 |
| hCoV-19/USA/IN-PU-LH000130674/2021 | EPI_ISL_1184199 |
| hCoV-19/USA/IN-PU-LH000130761/2021 | EPI_ISL_1184200 |
| hCoV-19/USA/IN-PU-LH000130882/2021 | EPI_ISL_1184195 |
| hCoV-19/USA/IN-PU-LH000125118/2021 | EPI_ISL_1184194 |
| hCoV-19/USA/IN-PU-LH000148197/2021 | EPI_ISL_1184196 |
| hCoV-19/USA/IN-PU-LH000128502/2021 | EPI_ISL_1184192 |
| hCoV-19/USA/IN-PU-LH000125086/2021 | EPI_ISL_1184191 |
| hCoV-19/USA/IN-PU-LH000130975/2021 | EPI_ISL_1184193 |
| hCoV-19/USA/IN-PU-LH000130952/2021 | EPI_ISL_1184197 |
| hCoV-19/USA/IN-PU-LH000125243/2021 | EPI_ISL_1184188 |
| hCoV-19/USA/IN-PU-LH000111675/2021 | EPI_ISL_1184187 |
| hCoV-19/USA/IN-PU-LH000125080/2021 | EPI_ISL_1184189 |
| hCoV-19/USA/IN-PU-LH000122049/2021 | EPI_ISL_1184185 |
| hCoV-19/USA/IN-PU-LH000122119/2021 | EPI_ISL_1184186 |
| hCoV-19/USA/IN-PU-LH000122390/2021 | EPI_ISL_1184190 |
| hCoV-19/USA/IN-PU-LH000125380/2021 | EPI_ISL_1184198 |
| hCoV-19/USA/IN-PU-LH000094305/2021 | EPI_ISL_1184201 |

| | |
|---|---|
| hCoV-19/USA/IN-PU-LH000141747/2021 | EPI_ISL_1184202 |
| hCoV-19/USA/IN-PU-LH000130743/2021 | EPI_ISL_1184211 |
| hCoV-19/USA/IN-PU-LH000137275/2021 | EPI_ISL_1184212 |
| hCoV-19/USA/IN-PU-LH000139428/2021 | EPI_ISL_1184214 |
| hCoV-19/USA/IN-PU-LH000137122/2021 | EPI_ISL_1184215 |
| hCoV-19/USA/IN-PU-LH000134305/2021 | EPI_ISL_1184219 |
| hCoV-19/USA/IN-PU-LH000137071/2021 | EPI_ISL_1184206 |
| hCoV-19/USA/IN-PU-LH000141747/2021 | EPI_ISL_1184202 |
| hCoV-19/USA/IN-PU-LH000130761/2021 | EPI_ISL_1184200 |
| hCoV-19/USA/IN-PU-LH000128502/2021 | EPI_ISL_1184192 |
| hCoV-19/USA/IN-PU-LH000122390/2021 | EPI_ISL_1184190 |
| hCoV-19/USA/IN-PU-LH000122119/2021 | EPI_ISL_1184186 |
| hCoV-19/USA/IN-PU-LH000122049/2021 | EPI_ISL_1184185 |
| hCoV-19/USA/IN-PU-LH000130952/2021 | EPI_ISL_1184197 |
| hCoV-19/USA/IN-PU-LH000145283/2021 | EPI_ISL_1184173 |
| hCoV-19/USA/IN-PU-LH000145457/2021 | EPI_ISL_1184171 |
| hCoV-19/USA/IN-PU-LH000125380/2021 | EPI_ISL_1184198 |
| hCoV-19/USA/IN-PU-LH000145467/2021 | EPI_ISL_1184172 |
| hCoV-19/USA/IN-PU-LH000146931/2021 | EPI_ISL_1255295 |
| hCoV-19/USA/IN-PU-LH000105391/2021 | EPI_ISL_1255294 |
| hCoV-19/USA/IN-PU-LH000144809/2021 | EPI_ISL_1255290 |
| hCoV-19/USA/IN-PU-LH000144791/2021 | EPI_ISL_1255289 |
| hCoV-19/USA/IN-PU-LH000146529/2021 | EPI_ISL_1255291 |
| hCoV-19/USA/IN-PU-LH000144713/2021 | EPI_ISL_1255287 |
| hCoV-19/USA/IN-PU-LH000144689/2021 | EPI_ISL_1255286 |
| hCoV-19/USA/IN-PU-LH000144762/2021 | EPI_ISL_1255288 |
| hCoV-19/USA/IN-PU-LH00043927/2021 | EPI_ISL_1255292 |

| | |
|---|---|
| hCoV-19/USA/IN-PU-LH000142608/2021 | EPI_ISL_1255283 |
| hCoV-19/USA/IN-PU-LH000137093/2021 | EPI_ISL_1255282 |
| hCoV-19/USA/IN-PU-LH000143108/2021 | EPI_ISL_1255284 |
| hCoV-19/USA/IN-PU-LH000129280/2021 | EPI_ISL_1255280 |
| hCoV-19/USA/IN-PU-LH000111714/2021 | EPI_ISL_1255279 |
| hCoV-19/USA/IN-PU-LH000134913/2021 | EPI_ISL_1255281 |
| hCoV-19/USA/IN-PU-LH000144595/2021 | EPI_ISL_1255285 |
| hCoV-19/USA/IN-PU-LH000084853/2021 | EPI_ISL_1255293 |
| hCoV-19/USA/IN-PU-LH000165434/2021 | EPI_ISL_1823477 |
| hCoV-19/USA/IN-PU-LH000161721/2021 | EPI_ISL_1823476 |
| hCoV-19/USA/IN-PU-LH000107081/2021 | EPI_ISL_1823441 |
| hCoV-19/USA/IN-PU-LH000136316/2021 | EPI_ISL_1823475 |
| hCoV-19/USA/IN-PU-LH000165181/2021 | EPI_ISL_1823474 |
| hCoV-19/USA/IN-PU-LH000167908/2021 | EPI_ISL_1823440 |
| hCoV-19/USA/IN-PU-LH000101815/2021 | EPI_ISL_1823420 |
| hCoV-19/USA/IN-PU-LH000136283/2021 | EPI_ISL_1823473 |
| hCoV-19/USA/IN-PU-LH000166576/2021 | EPI_ISL_1823472 |
| hCoV-19/USA/IN-PU-LH000111633/2021 | EPI_ISL_1823439 |
| hCoV-19/USA/IN-PU-LH000161602/2021 | EPI_ISL_1823471 |
| hCoV-19/USA/IN-PU-LH000111568/2021 | EPI_ISL_1823470 |
| hCoV-19/USA/IN-PU-LH000160461/2021 | EPI_ISL_1823437 |
| hCoV-19/USA/IN-PU-LH000101989/2021 | EPI_ISL_1823419 |
| hCoV-19/USA/IN-PU-LH000146820/2021 | EPI_ISL_1823410 |
| hCoV-19/USA/IN-PU-LH000161630/2021 | EPI_ISL_1823469 |
| hCoV-19/USA/IN-PU-LH000161906/2021 | EPI_ISL_1823468 |
| hCoV-19/USA/IN-PU-LH000111927/2021 | EPI_ISL_1823435 |
| hCoV-19/USA/IN-PU-LH000171261/2021 | EPI_ISL_1823525 |

| | |
|---|---|
| hCoV-19/USA/IN-PU-LH000166844/2021 | EPI_ISL_1823524 |
| hCoV-19/USA/IN-PU-LH000166580/2021 | EPI_ISL_1823465 |
| hCoV-19/USA/IN-PU-LH000166803/2021 | EPI_ISL_1823523 |
| hCoV-19/USA/IN-PU-LH000190793/2021 | EPI_ISL_1823526 |
| hCoV-19/USA/IN-PU-LH000190839/2021 | EPI_ISL_1823527 |
| hCoV-19/USA/IN-PU-LH000162853/2021 | EPI_ISL_1823528 |
| hCoV-19/USA/IN-PU-LH000161525/2021 | EPI_ISL_1823530 |
| hCoV-19/USA/IN-PU-LH000164253/2021 | EPI_ISL_1823531 |
| hCoV-19/USA/IN-PU-LH000166801/2021 | EPI_ISL_1823533 |
| hCoV-19/USA/IN-PU-LH000165357/2021 | EPI_ISL_1823534 |
| hCoV-19/USA/IN-PU-LH000190767/2021 | EPI_ISL_1823535 |
| hCoV-19/USA/IN-PU-LH000178976/2021 | EPI_ISL_1823536 |
| hCoV-19/USA/IN-PU-LH000146817/2021 | EPI_ISL_1823537 |
| hCoV-19/USA/IN-PU-LH000190900/2021 | EPI_ISL_1823538 |
| hCoV-19/USA/IN-PU-LH000175533/2021 | EPI_ISL_1823540 |
| hCoV-19/USA/IN-PU-LH000174029/2021 | EPI_ISL_1823541 |
| hCoV-19/USA/IN-PU-LH000174011/2021 | EPI_ISL_1823542 |
| hCoV-19/USA/IN-PU-LH000189923/2021 | EPI_ISL_1823543 |
| hCoV-19/USA/IN-PU-LH000153927/2021 | EPI_ISL_1823544 |
| hCoV-19/USA/IN-PU-LH000177024/2021 | EPI_ISL_1823545 |
| hCoV-19/USA/IN-PU-LH000179039/2021 | EPI_ISL_1823546 |
| hCoV-19/USA/IN-PU-LH000189506/2021 | EPI_ISL_1823547 |
| hCoV-19/USA/IN-PU-LH000189379/2021 | EPI_ISL_1823548 |
| hCoV-19/USA/IN-PU-LH000166910/2021 | EPI_ISL_1823522 |
| hCoV-19/USA/IN-PU-LH000165869/2021 | EPI_ISL_1823463 |
| hCoV-19/USA/IN-PU-LH000128349/2021 | EPI_ISL_1823434 |
| hCoV-19/USA/IN-PU-LH000105605/2021 | EPI_ISL_1823417 |

| | |
|---|---|
| hCoV-19/USA/IN-PU-LH000166874/2021 | EPI_ISL_1823521 |
| hCoV-19/USA/IN-PU-LH000166829/2021 | EPI_ISL_1823520 |
| hCoV-19/USA/IN-PU-LH000161674/2021 | EPI_ISL_1823462 |
| hCoV-19/USA/IN-PU-LH000166614/2021 | EPI_ISL_1823519 |
| hCoV-19/USA/IN-PU-LH000166714/2021 | EPI_ISL_1823518 |
| hCoV-19/USA/IN-PU-LH000161937/2021 | EPI_ISL_1823461 |
| hCoV-19/USA/IN-PU-LH000131612/2021 | EPI_ISL_1823433 |
| hCoV-19/USA/IN-PU-LH000161614/2021 | EPI_ISL_1823517 |
| hCoV-19/USA/IN-PU-LH000166655/2021 | EPI_ISL_1823516 |
| hCoV-19/USA/IN-PU-LH000161593/2021 | EPI_ISL_1823460 |
| hCoV-19/USA/IN-PU-LH000157633/2021 | EPI_ISL_1823515 |
| hCoV-19/USA/IN-PU-LH000162518/2021 | EPI_ISL_1823514 |
| hCoV-19/USA/IN-PU-LH000154511/2021 | EPI_ISL_1823459 |
| hCoV-19/USA/IN-PU-LH000138689/2021 | EPI_ISL_1823430 |
| hCoV-19/USA/IN-PU-LH000112925/2021 | EPI_ISL_1823416 |
| hCoV-19/USA/IN-PU-LH000146963/2021 | EPI_ISL_1823409 |
| hCoV-19/USA/IN-PU-LH000146703/2021 | EPI_ISL_1823406 |
| hCoV-19/USA/IN-PU-LH000152549/2021 | EPI_ISL_1823513 |
| hCoV-19/USA/IN-PU-LH000166549/2021 | EPI_ISL_1823512 |
| hCoV-19/USA/IN-PU-LH000161813/2021 | EPI_ISL_1823458 |
| hCoV-19/USA/IN-PU-LH000151004/2021 | EPI_ISL_1823457 |
| hCoV-19/USA/IN-PU-LH000190836/2021 | EPI_ISL_1823510 |
| hCoV-19/USA/IN-PU-LH000128259/2021 | EPI_ISL_1823428 |
| hCoV-19/USA/IN-PU-LH000189386/2021 | EPI_ISL_1823508 |
| hCoV-19/USA/IN-PU-LH000165201/2021 | EPI_ISL_1823456 |
| hCoV-19/USA/IN-PU-LH000190775/2021 | EPI_ISL_1823507 |
| hCoV-19/USA/IN-PU-LH000190787/2021 | EPI_ISL_1823506 |

| | |
|---|---|
| hCoV-19/USA/IN-PU-LH000165455/2021 | EPI_ISL_1823454 |
| hCoV-19/USA/IN-PU-LH000190899/2021 | EPI_ISL_1823505 |
| hCoV-19/USA/IN-PU-LH000101797/2021 | EPI_ISL_1823427 |
| hCoV-19/USA/IN-PU-LH000190893/2021 | EPI_ISL_1823504 |
| hCoV-19/USA/IN-PU-LH000165119/2021 | EPI_ISL_1823453 |
| hCoV-19/USA/IN-PU-LH000166879/2021 | EPI_ISL_1823503 |
| hCoV-19/USA/IN-PU-LH000152878/2021 | EPI_ISL_1823502 |
| hCoV-19/USA/IN-PU-LH000136098/2021 | EPI_ISL_1823452 |
| hCoV-19/USA/IN-PU-LH000089203/2021 | EPI_ISL_1823501 |
| hCoV-19/USA/IN-PU-LH000147463/2021 | EPI_ISL_1823426 |
| hCoV-19/USA/IN-PU-LH000161638/2021 | EPI_ISL_1823467 |
| hCoV-19/USA/IN-PU-LH000172706/2021 | EPI_ISL_1823500 |
| hCoV-19/USA/IN-PU-LH000190876/2021 | EPI_ISL_1823539 |
| hCoV-19/USA/IN-PU-LH000161862/2021 | EPI_ISL_1823481 |
| hCoV-19/USA/IN-PU-LH000126879/2021 | EPI_ISL_1823451 |
| hCoV-19/USA/IN-PU-LH000166894/2021 | EPI_ISL_1823499 |
| hCoV-19/USA/IN-PU-LH000166665/2021 | EPI_ISL_1823466 |
| hCoV-19/USA/IN-PU-LH000152631/2021 | EPI_ISL_1823529 |
| hCoV-19/USA/IN-PU-LH000105534/2021 | EPI_ISL_1823414 |
| hCoV-19/USA/IN-PU-LH000166650/2021 | EPI_ISL_1823498 |
| hCoV-19/USA/IN-PU-LH000111892/2021 | EPI_ISL_1823415 |
| hCoV-19/USA/IN-PU-LH000111761/2021 | EPI_ISL_1823408 |
| hCoV-19/USA/IN-PU-LH000165133/2021 | EPI_ISL_1823450 |
| hCoV-19/USA/IN-PU-LH000166713/2021 | EPI_ISL_1823497 |
| hCoV-19/USA/IN-PU-LH000111959/2021 | EPI_ISL_1823425 |
| hCoV-19/USA/IN-PU-LH000140648/2021 | EPI_ISL_1823438 |
| hCoV-19/USA/IN-PU-LH000043798/2021 | EPI_ISL_1823532 |

| | |
|---|---|
| hCoV-19/USA/IN-PU-LH000111624/2021 | EPI_ISL_1823405 |
| hCoV-19/USA/IN-PU-LH000165387/2021 | EPI_ISL_1823496 |
| hCoV-19/USA/IN-PU-LH000101558/2021 | EPI_ISL_1823404 |
| hCoV-19/USA/IN-PU-LH000111914/2021 | EPI_ISL_1823449 |
| hCoV-19/USA/IN-PU-LH000166636/2021 | EPI_ISL_1823495 |
| hCoV-19/USA/IN-PU-LH000146858/2021 | EPI_ISL_1823407 |
| hCoV-19/USA/IN-PU-LH000101777/2021 | EPI_ISL_1823418 |
| hCoV-19/USA/IN-PU-LH000111777/2021 | EPI_ISL_1823448 |
| hCoV-19/USA/IN-PU-LH000166632/2021 | EPI_ISL_1823493 |
| hCoV-19/USA/IN-PU-LH000112925/2021 | EPI_ISL_1823416 |
| hCoV-19/USA/IN-PU-LH000081212/2021 | EPI_ISL_1823424 |
| hCoV-19/USA/IN-PU-LH000176724/2021 | EPI_ISL_1823509 |
| hCoV-19/USA/IN-PU-LH000166616/2021 | EPI_ISL_1823494 |
| hCoV-19/USA/IN-PU-LH000166615/2021 | EPI_ISL_1823492 |
| hCoV-19/USA/IN-PU-LH000111579/2021 | EPI_ISL_1823413 |
| hCoV-19/USA/IN-PU-LH000128233/2021 | EPI_ISL_1823431 |
| hCoV-19/USA/IN-PU-LH000166579/2021 | EPI_ISL_1823489 |
| hCoV-19/USA/IN-PU-LH000146891/2021 | EPI_ISL_1823566 |
| hCoV-19/USA/IN-PU-LH000174057/2021 | EPI_ISL_1823565 |
| hCoV-19/USA/IN-PU-LH000166544/2021 | EPI_ISL_1823487 |
| hCoV-19/USA/IN-PU-LH000167635/2021 | EPI_ISL_1823446 |
| hCoV-19/USA/IN-PU-LH000101873/2021 | EPI_ISL_1823423 |
| hCoV-19/USA/IN-PU-LH000190876/2021 | EPI_ISL_1823539 |
| hCoV-19/USA/IN-PU-LH000146837/2021 | EPI_ISL_1823564 |
| hCoV-19/USA/IN-PU-LH000190831/2021 | EPI_ISL_1823563 |
| hCoV-19/USA/IN-PU-LH000105605/2021 | EPI_ISL_1823417 |
| hCoV-19/USA/IN-PU-LH000161932/2021 | EPI_ISL_1823486 |

| | |
|---|---|
| hCoV-19/USA/IN-PU-LH000173055/2021 | EPI_ISL_1823562 |
| hCoV-19/USA/IN-PU-LH000151660/2021 | EPI_ISL_1823561 |
| hCoV-19/USA/IN-PU-LH000161534/2021 | EPI_ISL_1823485 |
| hCoV-19/USA/IN-PU-LH000158494/2021 | EPI_ISL_1823445 |
| hCoV-19/USA/IN-PU-LH000097312/2021 | EPI_ISL_1823560 |
| hCoV-19/USA/IN-PU-LH000190754/2021 | EPI_ISL_1823559 |
| hCoV-19/USA/IN-PU-LH000155452/2021 | EPI_ISL_1823484 |
| hCoV-19/USA/IN-PU-LH000175511/2021 | EPI_ISL_1823558 |
| hCoV-19/USA/IN-PU-LH000190882/2021 | EPI_ISL_1823557 |
| hCoV-19/USA/IN-PU-LH000165687/2021 | EPI_ISL_1823483 |
| hCoV-19/USA/IN-PU-LH000165484/2021 | EPI_ISL_1823444 |
| hCoV-19/USA/IN-PU-LH000084696/2021 | EPI_ISL_1823422 |
| hCoV-19/USA/IN-PU-LH000189404/2021 | EPI_ISL_1823556 |
| hCoV-19/USA/IN-PU-LH000174093/2021 | EPI_ISL_1823555 |
| hCoV-19/USA/IN-PU-LH000101989/2021 | EPI_ISL_1823419 |
| hCoV-19/USA/IN-PU-LH000161809/2021 | EPI_ISL_1823482 |
| hCoV-19/USA/IN-PU-LH000097548/2021 | EPI_ISL_1823554 |
| hCoV-19/USA/IN-PU-LH000043629/2021 | EPI_ISL_1823553 |
| hCoV-19/USA/IN-PU-LH000161782/2021 | EPI_ISL_1823480 |
| hCoV-19/USA/IN-PU-LH000128156/2021 | EPI_ISL_1823443 |
| hCoV-19/USA/IN-PU-LH000174037/2021 | EPI_ISL_1823552 |
| hCoV-19/USA/IN-PU-LH000166934/2021 | EPI_ISL_1823551 |
| hCoV-19/USA/IN-PU-LH000111964/2021 | EPI_ISL_1823479 |
| hCoV-19/USA/IN-PU-LH000164236/2021 | EPI_ISL_1823550 |
| hCoV-19/USA/IN-PU-LH000189330/2021 | EPI_ISL_1823549 |
| hCoV-19/USA/IN-PU-LH000161725/2021 | EPI_ISL_1823478 |
| hCoV-19/USA/IN-PU-LH000128227/2021 | EPI_ISL_1823442 |

| | |
|---|---|
| hCoV-19/USA/IN-PU-LH000101997/2021 | EPI_ISL_1823421 |
| hCoV-19/USA/IN-PU-LH000105428/2021 | EPI_ISL_1823411 |
| hCoV-19/USA/IN-PU-LH000174011/2021 | EPI_ISL_1823542 |
| hCoV-19/USA/IN-PU-LH000101777/2021 | EPI_ISL_1823418 |
| hCoV-19/USA/IN-PU-LH000128233/2021 | EPI_ISL_1823431 |
| hCoV-19/USA/IN-PU-LH000128375/2021 | EPI_ISL_1823432 |
| hCoV-19/USA/IN-PU-LH000165084/2021 | EPI_ISL_1823436 |
| hCoV-19/USA/IN-PU-LH000140648/2021 | EPI_ISL_1823438 |
| hCoV-19/USA/IN-PU-LH000166665/2021 | EPI_ISL_1823466 |
| hCoV-19/USA/IN-PU-LH000161638/2021 | EPI_ISL_1823467 |
| hCoV-19/USA/IN-PU-LH000189685/2021 | EPI_ISL_2304460 |
| hCoV-19/USA/IN-PU-LH000177872/2021 | EPI_ISL_2304458 |
| hCoV-19/USA/IN-PU-LH000115645/2021 | EPI_ISL_2304457 |
| hCoV-19/USA/IN-PU-LH000179466/2021 | EPI_ISL_2304453 |
| hCoV-19/USA/IN-PU-LH000189756/2021 | EPI_ISL_2304451 |
| hCoV-19/USA/IN-PU-LH000183890/2021 | EPI_ISL_2304449 |
| hCoV-19/USA/IN-PU-LH000189737/2021 | EPI_ISL_2304446 |
| hCoV-19/USA/IN-PU-LH000187551/2021 | EPI_ISL_2304444 |
| hCoV-19/USA/IN-PU-LH000171133/2021 | EPI_ISL_2304442 |
| hCoV-19/USA/IN-PU-LH000159133/2021 | EPI_ISL_2304441 |
| hCoV-19/USA/IN-PU-LH000184440/2021 | EPI_ISL_2304439 |
| hCoV-19/USA/IN-PU-LH000183994/2021 | EPI_ISL_2304438 |
| hCoV-19/USA/IN-PU-LH000183853/2021 | EPI_ISL_2304437 |
| hCoV-19/USA/IN-PU-LH000190170/2021 | EPI_ISL_2304436 |
| hCoV-19/USA/IN-PU-LH000179679/2021 | EPI_ISL_2304433 |
| hCoV-19/USA/IN-PU-LH000180976/2021 | EPI_ISL_2304432 |
| hCoV-19/USA/IN-PU-LH000185841/2021 | EPI_ISL_2304431 |

| | |
|---|---|
| hCoV-19/USA/IN-PU-LH000172626/2021 | EPI_ISL_2304430 |
| hCoV-19/USA/IN-PU-LH000181970/2021 | EPI_ISL_2304428 |
| hCoV-19/USA/IN-PU-LH000176624/2021 | EPI_ISL_2304427 |
| hCoV-19/USA/IN-PU-LH000187880/2021 | EPI_ISL_2304426 |
| hCoV-19/USA/IN-PU-LH000171098/2021 | EPI_ISL_2304425 |
| hCoV-19/USA/IN-PU-LH000171138/2021 | EPI_ISL_2304424 |
| hCoV-19/USA/IN-PU-LH000184489/2021 | EPI_ISL_2304423 |
| hCoV-19/USA/IN-PU-LH000183886/2021 | EPI_ISL_2304422 |
| hCoV-19/USA/IN-PU-LH000180698/2021 | EPI_ISL_2304421 |
| hCoV-19/USA/IN-PU-LH000183590/2021 | EPI_ISL_2304420 |
| hCoV-19/USA/IN-PU-LH000188675/2021 | EPI_ISL_2304417 |
| hCoV-19/USA/IN-PU-LH000175112/2021 | EPI_ISL_2304416 |
| hCoV-19/USA/IN-PU-LH000177043/2021 | EPI_ISL_2304415 |
| hCoV-19/USA/IN-PU-LH000179622/2021 | EPI_ISL_2304414 |
| hCoV-19/USA/IN-PU-LH000104681/2021 | EPI_ISL_2304413 |
| hCoV-19/USA/IN-PU-LH000186929/2021 | EPI_ISL_2304412 |
| hCoV-19/USA/IN-PU-LH000171088/2021 | EPI_ISL_2304411 |
| hCoV-19/USA/IN-PU-LH000176477/2021 | EPI_ISL_2304409 |
| hCoV-19/USA/IN-PU-LH000176693/2021 | EPI_ISL_2304407 |
| hCoV-19/USA/IN-PU-LH000176380/2021 | EPI_ISL_2304406 |
| hCoV-19/USA/IN-PU-LH000190871/2021 | EPI_ISL_2304405 |
| hCoV-19/USA/IN-PU-LH000115808/2021 | EPI_ISL_2304404 |
| hCoV-19/USA/IN-PU-LH000171082/2021 | EPI_ISL_2304403 |
| hCoV-19/USA/IN-PU-LH000187287/2021 | EPI_ISL_2304402 |
| hCoV-19/USA/IN-PU-LH000188025/2021 | EPI_ISL_2304401 |
| hCoV-19/USA/IN-PU-LH000188136/2021 | EPI_ISL_2304400 |
| hCoV-19/USA/IN-PU-LH000171159/2021 | EPI_ISL_2304399 |

| | |
|---|---|
| hCoV-19/USA/IN-PU-LH000173402/2021 | EPI_ISL_2304398 |
| hCoV-19/USA/IN-PU-LH000181484/2021 | EPI_ISL_2304396 |
| hCoV-19/USA/IN-PU-LH000174716/2021 | EPI_ISL_2304394 |
| hCoV-19/USA/IN-PU-LH000172533/2021 | EPI_ISL_2304393 |
| hCoV-19/USA/IN-PU-LH000181807/2021 | EPI_ISL_2304392 |
| hCoV-19/USA/IN-PU-LH000180366/2021 | EPI_ISL_2304391 |
| hCoV-19/USA/IN-PU-LH000182994/2021 | EPI_ISL_2304390 |
| hCoV-19/USA/IN-PU-LH000181359/2021 | EPI_ISL_2304389 |
| hCoV-19/USA/IN-PU-LH000175112/2021 | EPI_ISL_2304416 |
| hCoV-19/USA/IN-PU-LH000177043/2021 | EPI_ISL_2304415 |
| hCoV-19/USA/IN-PU-LH000179622/2021 | EPI_ISL_2304414 |
| hCoV-19/USA/IN-PU-LH000104681/2021 | EPI_ISL_2304413 |
| hCoV-19/USA/IN-PU-LH000186929/2021 | EPI_ISL_2304412 |
| hCoV-19/USA/IN-PU-LH000171088/2021 | EPI_ISL_2304411 |
| hCoV-19/USA/IN-PU-LH000176477/2021 | EPI_ISL_2304409 |
| hCoV-19/USA/IN-PU-LH000176693/2021 | EPI_ISL_2304407 |
| hCoV-19/USA/IN-PU-LH000176380/2021 | EPI_ISL_2304406 |
| hCoV-19/USA/IN-PU-LH000190871/2021 | EPI_ISL_2304405 |
| hCoV-19/USA/IN-PU-LH000115808/2021 | EPI_ISL_2304404 |
| hCoV-19/USA/IN-PU-LH000171082/2021 | EPI_ISL_2304403 |
| hCoV-19/USA/IN-PU-LH000187287/2021 | EPI_ISL_2304402 |
| hCoV-19/USA/IN-PU-LH000188025/2021 | EPI_ISL_2304401 |
| hCoV-19/USA/IN-PU-LH000188136/2021 | EPI_ISL_2304400 |
| hCoV-19/USA/IN-PU-LH000171159/2021 | EPI_ISL_2304399 |
| hCoV-19/USA/IN-PU-LH000173402/2021 | EPI_ISL_2304398 |
| hCoV-19/USA/IN-PU-LH000181484/2021 | EPI_ISL_2304396 |
| hCoV-19/USA/IN-PU-LH000174716/2021 | EPI_ISL_2304394 |

| | |
|---|---|
| hCoV-19/USA/IN-PU-LH000172533/2021 | EPI_ISL_2304393 |
| hCoV-19/USA/IN-PU-LH000181807/2021 | EPI_ISL_2304392 |
| hCoV-19/USA/IN-PU-LH000180366/2021 | EPI_ISL_2304391 |
| hCoV-19/USA/IN-PU-LH000182994/2021 | EPI_ISL_2304390 |
| hCoV-19/USA/IN-PU-LH000181359/2021 | EPI_ISL_2304389 |
| hCoV-19/USA/IN-PU-LH000177963/2021 | EPI_ISL_2304388 |
| hCoV-19/USA/IN-PU-LH000181983/2021 | EPI_ISL_2304387 |
| hCoV-19/USA/IN-PU-LH000185305/2021 | EPI_ISL_2304386 |
| hCoV-19/USA/IN-PU-LH000185162/2021 | EPI_ISL_2304385 |
| hCoV-19/USA/IN-PU-LH000176641/2021 | EPI_ISL_2304384 |
| hCoV-19/USA/IN-PU-LH000171767/2021 | EPI_ISL_2304383 |
| hCoV-19/USA/IN-PU-LH000187686/2021 | EPI_ISL_2304382 |
| hCoV-19/USA/IN-PU-LH000176634/2021 | EPI_ISL_2304381 |
| hCoV-19/USA/IN-PU-LH000176658/2021 | EPI_ISL_2304380 |
| hCoV-19/USA/IN-PU-LH000174005/2021 | EPI_ISL_2304379 |
| hCoV-19/USA/IN-PU-LH000181377/2021 | EPI_ISL_2304378 |
| hCoV-19/USA/IN-PU-LH000176711/2021 | EPI_ISL_2304377 |
| hCoV-19/USA/IN-PU-LH000173139/2021 | EPI_ISL_2304373 |
| hCoV-19/USA/IN-PU-LH000175466/2021 | EPI_ISL_2304372 |
| hCoV-19/USA/IN-PU-LH000172545/2021 | EPI_ISL_2304371 |
| hCoV-19/USA/IN-PU-LH000143627/2021 | EPI_ISL_2304370 |
| hCoV-19/USA/IN-PU-LH000191319/2021 | EPI_ISL_2304369 |
| hCoV-19/USA/IN-PU-LH000171970/2021 | EPI_ISL_2304368 |
| hCoV-19/USA/IN-PU-LH000179679/2021 | EPI_ISL_2304433 |
| hCoV-19/USA/IN-PU-LH000190170/2021 | EPI_ISL_2304436 |
| hCoV-19/USA/IN-PU-LH000183853/2021 | EPI_ISL_2304437 |
| hCoV-19/USA/IN-PU-LH000183994/2021 | EPI_ISL_2304438 |

| | |
|---|---|
| hCoV-19/USA/IN-PU-LH000184440/2021 | EPI_ISL_2304439 |
| hCoV-19/USA/IN-PU-LH000159133/2021 | EPI_ISL_2304441 |
| hCoV-19/USA/IN-PU-LH000171133/2021 | EPI_ISL_2304442 |
| hCoV-19/USA/IN-PU-LH000187551/2021 | EPI_ISL_2304444 |
| hCoV-19/USA/IN-PU-LH000189737/2021 | EPI_ISL_2304446 |
| hCoV-19/USA/IN-PU-LH000183890/2021 | EPI_ISL_2304449 |
| hCoV-19/USA/IN-PU-LH000189756/2021 | EPI_ISL_2304451 |
| hCoV-19/USA/IN-PU-LH000179466/2021 | EPI_ISL_2304453 |
| hCoV-19/USA/IN-PU-LH000115645/2021 | EPI_ISL_2304457 |
| hCoV-19/USA/IN-PU-LH000177872/2021 | EPI_ISL_2304458 |
| hCoV-19/USA/IN-PU-LH000189685/2021 | EPI_ISL_2304460 |
| hCoV-19/USA/IN-PU-LH000187547/2021 | EPI_ISL_2304443 |
| hCoV-19/USA/IN-PU-LH000189891/2021 | EPI_ISL_2304440 |
| hCoV-19/USA/IN-PU-LH000115704/2021 | EPI_ISL_2304445 |
| hCoV-19/USA/IN-PU-LH000189787/2021 | EPI_ISL_2304447 |
| hCoV-19/USA/IN-PU-LH000183907/2021 | EPI_ISL_2304448 |
| hCoV-19/USA/IN-PU-LH000185993/2021 | EPI_ISL_2304450 |
| hCoV-19/USA/IN-PU-lh000082308/2021 | EPI_ISL_5990416 |
| hCoV-19/USA/IN-PU-lh000111788/2021 | EPI_ISL_5990219 |
| hCoV-19/USA/IN-PU-lh000115424/2021 | EPI_ISL_5990430 |
| hCoV-19/USA/IN-PU-lh000043557/2021 | EPI_ISL_5990206 |
| hCoV-19/USA/IN-PU-lh000094033/2021 | EPI_ISL_5990207 |
| hCoV-19/USA/IN-PU-lh000094088/2021 | EPI_ISL_5990208 |
| hCoV-19/USA/IN-PU-lh000132517/2021 | EPI_ISL_5990209 |
| hCoV-19/USA/IN-PU-lh000132574/2021 | EPI_ISL_5990210 |
| hCoV-19/USA/IN-PU-lh000100073/2021 | EPI_ISL_5990211 |
| hCoV-19/USA/IN-PU-lh000132796/2021 | EPI_ISL_5990212 |

| | |
|---|---|
| hCoV-19/USA/IN-PU-lh000121532/2021 | EPI_ISL_5990213 |
| hCoV-19/USA/IN-PU-lh000132742/2021 | EPI_ISL_5990214 |
| hCoV-19/USA/IN-PU-lh000094335/2021 | EPI_ISL_5990215 |
| hCoV-19/USA/IN-PU-lh000121800/2021 | EPI_ISL_5990216 |
| hCoV-19/USA/IN-PU-lh000121857/2021 | EPI_ISL_5990217 |
| hCoV-19/USA/IN-PU-lh000121864/2021 | EPI_ISL_5990218 |
| hCoV-19/USA/IN-PU-lh000111702/2021 | EPI_ISL_5990220 |
| hCoV-19/USA/IN-PU-lh000119980/2021 | EPI_ISL_5990221 |
| hCoV-19/USA/IN-PU-lh000106554/2021 | EPI_ISL_5990222 |
| hCoV-19/USA/IN-PU-lh000100219/2021 | EPI_ISL_5990223 |
| hCoV-19/USA/IN-PU-lh000121801/2021 | EPI_ISL_5990224 |
| hCoV-19/USA/IN-PU-lh000111763/2021 | EPI_ISL_5990225 |
| hCoV-19/USA/IN-PU-lh000121894/2021 | EPI_ISL_5990226 |
| hCoV-19/USA/IN-PU-lh000111882/2021 | EPI_ISL_5990227 |
| hCoV-19/USA/IN-PU-lh000121826/2021 | EPI_ISL_5990228 |
| hCoV-19/USA/IN-PU-lh000121758/2021 | EPI_ISL_5990229 |
| hCoV-19/USA/IN-PU-lh000125694/2021 | EPI_ISL_5990230 |
| hCoV-19/USA/IN-PU-lh000121882/2021 | EPI_ISL_5990231 |
| hCoV-19/USA/IN-PU-lh000122009/2021 | EPI_ISL_5990232 |
| hCoV-19/USA/IN-PU-lh000122422/2021 | EPI_ISL_5990233 |
| hCoV-19/USA/IN-PU-lh000130986/2021 | EPI_ISL_5990234 |
| hCoV-19/USA/IN-PU-lh000121542/2021 | EPI_ISL_5990235 |
| hCoV-19/USA/IN-PU-lh000121583/2021 | EPI_ISL_5990236 |
| hCoV-19/USA/IN-PU-lh000130928/2021 | EPI_ISL_5990237 |
| hCoV-19/USA/IN-PU-lh000130974/2021 | EPI_ISL_5990238 |
| hCoV-19/USA/IN-PU-lh000137182/2021 | EPI_ISL_5990239 |
| hCoV-19/USA/IN-PU-lh000137512/2021 | EPI_ISL_5990240 |

| | |
|---|---|
| hCoV-19/USA/IN-PU-lh000137170/2021 | EPI_ISL_5990241 |
| hCoV-19/USA/IN-PU-lh000141676/2021 | EPI_ISL_5990242 |
| hCoV-19/USA/IN-PU-lh000122223/2021 | EPI_ISL_5990243 |
| hCoV-19/USA/IN-PU-lh000125333/2021 | EPI_ISL_5990244 |
| hCoV-19/USA/IN-PU-lh000137166/2021 | EPI_ISL_5990245 |
| hCoV-19/USA/IN-PU-lh000111566/2021 | EPI_ISL_5990246 |
| hCoV-19/USA/IN-PU-lh000137325/2021 | EPI_ISL_5990247 |
| hCoV-19/USA/IN-PU-lh000137155/2021 | EPI_ISL_5990248 |
| hCoV-19/USA/IN-PU-lh000133519/2021 | EPI_ISL_5990249 |
| hCoV-19/USA/IN-PU-lh000134450/2021 | EPI_ISL_5990250 |
| hCoV-19/USA/IN-PU-lh000144527/2021 | EPI_ISL_5990251 |
| hCoV-19/USA/IN-PU-lh000137271/2021 | EPI_ISL_5990252 |
| hCoV-19/USA/IN-PU-lh000137445/2021 | EPI_ISL_5990253 |
| hCoV-19/USA/IN-PU-lh000137439/2021 | EPI_ISL_5990254 |
| hCoV-19/USA/IN-PU-lh000137209/2021 | EPI_ISL_5990255 |
| hCoV-19/USA/IN-PU-lh000144566/2021 | EPI_ISL_5990256 |
| hCoV-19/USA/IN-PU-lh000144547/2021 | EPI_ISL_5990257 |
| hCoV-19/USA/IN-PU-lh000144862/2021 | EPI_ISL_5990258 |
| hCoV-19/USA/IN-PU-lh000144786/2021 | EPI_ISL_5990259 |
| hCoV-19/USA/IN-PU-lh000144665/2021 | EPI_ISL_5990260 |
| hCoV-19/USA/IN-PU-lh000144663/2021 | EPI_ISL_5990261 |
| hCoV-19/USA/IN-PU-lh000146503/2021 | EPI_ISL_5990262 |
| hCoV-19/USA/IN-PU-lh000144748/2021 | EPI_ISL_5990263 |
| hCoV-19/USA/IN-PU-lh000111522/2021 | EPI_ISL_5990264 |
| hCoV-19/USA/IN-PU-lh000129209/2021 | EPI_ISL_5990265 |
| hCoV-19/USA/IN-PU-lh000144912/2021 | EPI_ISL_5990266 |
| hCoV-19/USA/IN-PU-lh000130924/2021 | EPI_ISL_5990267 |

| | |
|---|---|
| hCoV-19/USA/IN-PU-lh000111891/2021 | EPI_ISL_5990268 |
| hCoV-19/USA/IN-PU-lh000130960/2021 | EPI_ISL_5990269 |
| hCoV-19/USA/IN-PU-lh000146818/2021 | EPI_ISL_5990270 |
| hCoV-19/USA/IN-PU-lh000146802/2021 | EPI_ISL_5990271 |
| hCoV-19/USA/IN-PU-lh000105385/2021 | EPI_ISL_5990272 |
| hCoV-19/USA/IN-PU-lh000105322/2021 | EPI_ISL_5990273 |
| hCoV-19/USA/IN-PU-lh000146992/2021 | EPI_ISL_5990274 |
| hCoV-19/USA/IN-PU-lh000105401/2021 | EPI_ISL_5990275 |
| hCoV-19/USA/IN-PU-lh000105526/2021 | EPI_ISL_5990276 |
| hCoV-19/USA/IN-PU-lh000146987/2021 | EPI_ISL_5990277 |
| hCoV-19/USA/IN-PU-lh000105772/2021 | EPI_ISL_5990278 |
| hCoV-19/USA/IN-PU-lh000105545/2021 | EPI_ISL_5990279 |
| hCoV-19/USA/IN-PU-lh000105651/2021 | EPI_ISL_5990280 |
| hCoV-19/USA/IN-PU-lh000105540/2021 | EPI_ISL_5990281 |
| hCoV-19/USA/IN-PU-lh000105696/2021 | EPI_ISL_5990282 |
| hCoV-19/USA/IN-PU-lh000105718/2021 | EPI_ISL_5990283 |
| hCoV-19/USA/IN-PU-lh000157105/2021 | EPI_ISL_5990284 |
| hCoV-19/USA/IN-PU-lh000146885/2021 | EPI_ISL_5990285 |
| hCoV-19/USA/IN-PU-lh000116202/2021 | EPI_ISL_5990286 |
| hCoV-19/USA/IN-PU-lh000116178/2021 | EPI_ISL_5990287 |
| hCoV-19/USA/IN-PU-lh000116500/2021 | EPI_ISL_5990288 |
| hCoV-19/USA/IN-PU-lh000116312/2021 | EPI_ISL_5990289 |
| hCoV-19/USA/IN-PU-lh000116256/2021 | EPI_ISL_5990290 |
| hCoV-19/USA/IN-PU-lh000116068/2021 | EPI_ISL_5990291 |
| hCoV-19/USA/IN-PU-lh000116177/2021 | EPI_ISL_5990292 |
| hCoV-19/USA/IN-PU-lh000116275/2021 | EPI_ISL_5990293 |
| hCoV-19/USA/IN-PU-lh000101188/2021 | EPI_ISL_5990294 |

| | |
|---|---|
| hCoV-19/USA/IN-PU-lh000116292/2021 | EPI_ISL_5990295 |
| hCoV-19/USA/IN-PU-lh000116185/2021 | EPI_ISL_5990296 |
| hCoV-19/USA/IN-PU-lh000116429/2021 | EPI_ISL_5990297 |
| hCoV-19/USA/IN-PU-lh000116108/2021 | EPI_ISL_5990298 |
| hCoV-19/USA/IN-PU-lh000116439/2021 | EPI_ISL_5990299 |
| hCoV-19/USA/IN-PU-lh000115264/2021 | EPI_ISL_5990300 |
| hCoV-19/USA/IN-PU-lh000010575/2021 | EPI_ISL_5990301 |
| hCoV-19/USA/IN-PU-lh000086980/2021 | EPI_ISL_5990302 |
| hCoV-19/USA/IN-PU-lh000086853/2021 | EPI_ISL_5990303 |
| hCoV-19/USA/IN-PU-lh000097818/2021 | EPI_ISL_5990304 |
| hCoV-19/USA/IN-PU-lh000086917/2021 | EPI_ISL_5990305 |
| hCoV-19/USA/IN-PU-lh000080511/2021 | EPI_ISL_5990306 |
| hCoV-19/USA/IN-PU-lh000080773/2021 | EPI_ISL_5990307 |
| hCoV-19/USA/IN-PU-lh000086858/2021 | EPI_ISL_5990308 |
| hCoV-19/USA/IN-PU-lh000080528/2021 | EPI_ISL_5990309 |
| hCoV-19/USA/IN-PU-lh000080594/2021 | EPI_ISL_5990310 |
| hCoV-19/USA/IN-PU-lh000086914/2021 | EPI_ISL_5990311 |
| hCoV-19/USA/IN-PU-lh000085105/2021 | EPI_ISL_5990312 |
| hCoV-19/USA/IN-PU-lh000080595/2021 | EPI_ISL_5990313 |
| hCoV-19/USA/IN-PU-lh000080772/2021 | EPI_ISL_5990314 |
| hCoV-19/USA/IN-PU-lh000145262/2021 | EPI_ISL_5990315 |
| hCoV-19/USA/IN-PU-lh000100586/2021 | EPI_ISL_5990316 |
| hCoV-19/USA/IN-PU-lh000082273/2021 | EPI_ISL_5990317 |
| hCoV-19/USA/IN-PU-lh000094106/2021 | EPI_ISL_5990318 |
| hCoV-19/USA/IN-PU-lh000082146/2021 | EPI_ISL_5990319 |
| hCoV-19/USA/IN-PU-lh000145143/2021 | EPI_ISL_5990320 |
| hCoV-19/USA/IN-PU-lh000145278/2021 | EPI_ISL_5990321 |

| | |
|---|---|
| hCoV-19/USA/IN-PU-lh000082246/2021 | EPI_ISL_5990322 |
| hCoV-19/USA/IN-PU-lh000094432/2021 | EPI_ISL_5990323 |
| hCoV-19/USA/IN-PU-lh000115244/2021 | EPI_ISL_5990324 |
| hCoV-19/USA/IN-PU-lh000094457/2021 | EPI_ISL_5990325 |
| hCoV-19/USA/IN-PU-lh000109445/2021 | EPI_ISL_5990326 |
| hCoV-19/USA/IN-PU-lh000109398/2021 | EPI_ISL_5990328 |
| hCoV-19/USA/IN-PU-lh000110887/2021 | EPI_ISL_5990329 |
| hCoV-19/USA/IN-PU-lh000100983/2021 | EPI_ISL_5990330 |
| hCoV-19/USA/IN-PU-lh000101161/2021 | EPI_ISL_5990331 |
| hCoV-19/USA/IN-PU-lh000116122/2021 | EPI_ISL_5990332 |
| hCoV-19/USA/IN-PU-lh000101347/2021 | EPI_ISL_5990333 |
| hCoV-19/USA/IN-PU-lh000101466/2021 | EPI_ISL_5990334 |
| hCoV-19/USA/IN-PU-lh000101068/2021 | EPI_ISL_5990335 |
| hCoV-19/USA/IN-PU-lh000094063/2021 | EPI_ISL_5990336 |
| hCoV-19/USA/IN-PU-lh000086865/2021 | EPI_ISL_5990337 |
| hCoV-19/USA/IN-PU-lh000094286/2021 | EPI_ISL_5990338 |
| hCoV-19/USA/IN-PU-lh000085958/2021 | EPI_ISL_5990339 |
| hCoV-19/USA/IN-PU-lh000094396/2021 | EPI_ISL_5990340 |
| hCoV-19/USA/IN-PU-lh000082633/2021 | EPI_ISL_5990341 |
| hCoV-19/USA/IN-PU-lh000082610/2021 | EPI_ISL_5990342 |
| hCoV-19/USA/IN-PU-lh000086764/2021 | EPI_ISL_5990343 |
| hCoV-19/USA/IN-PU-lh000080656/2021 | EPI_ISL_5990344 |
| hCoV-19/USA/IN-PU-lh000086872/2021 | EPI_ISL_5990345 |
| hCoV-19/USA/IN-PU-lh000086755/2021 | EPI_ISL_5990346 |
| hCoV-19/USA/IN-PU-lh000080581/2021 | EPI_ISL_5990347 |
| hCoV-19/USA/IN-PU-lh000080584/2021 | EPI_ISL_5990348 |
| hCoV-19/USA/IN-PU-lh000080776/2021 | EPI_ISL_5990349 |

| | |
|---|---|
| hCoV-19/USA/IN-PU-lh000080718/2021 | EPI_ISL_5990350 |
| hCoV-19/USA/IN-PU-lh000080714/2021 | EPI_ISL_5990351 |
| hCoV-19/USA/IN-PU-lh000104035/2021 | EPI_ISL_5990352 |
| hCoV-19/USA/IN-PU-lh000080706/2021 | EPI_ISL_5990353 |
| hCoV-19/USA/IN-PU-lh000080848/2021 | EPI_ISL_5990354 |
| hCoV-19/USA/IN-PU-lh000080572/2021 | EPI_ISL_5990355 |
| hCoV-19/USA/IN-PU-lh000086920/2021 | EPI_ISL_5990356 |
| hCoV-19/USA/IN-PU-lh000124668/2021 | EPI_ISL_5990357 |
| hCoV-19/USA/IN-PU-lh000109122/2021 | EPI_ISL_5990358 |
| hCoV-19/USA/IN-PU-lh000109009/2021 | EPI_ISL_5990359 |
| hCoV-19/USA/IN-PU-lh000130880/2021 | EPI_ISL_5990360 |
| hCoV-19/USA/IN-PU-lh000111561/2021 | EPI_ISL_5990361 |
| hCoV-19/USA/IN-PU-lh000105463/2021 | EPI_ISL_5990362 |
| hCoV-19/USA/IN-PU-lh000130658/2021 | EPI_ISL_5990363 |
| hCoV-19/USA/IN-PU-lh000137287/2021 | EPI_ISL_5990364 |
| hCoV-19/USA/IN-PU-lh000137119/2021 | EPI_ISL_5990365 |
| hCoV-19/USA/IN-PU-lh000084676/2021 | EPI_ISL_5990366 |
| hCoV-19/USA/IN-PU-lh000063970/2021 | EPI_ISL_5990367 |
| hCoV-19/USA/IN-PU-lh000132476/2021 | EPI_ISL_5990368 |
| hCoV-19/USA/IN-PU-lh000109024/2021 | EPI_ISL_5990369 |
| hCoV-19/USA/IN-PU-lh000101708/2021 | EPI_ISL_5990370 |
| hCoV-19/USA/IN-PU-lh000156554/2021 | EPI_ISL_5990371 |
| hCoV-19/USA/IN-PU-lh000104097/2021 | EPI_ISL_5990372 |
| hCoV-19/USA/IN-PU-lh000080520/2021 | EPI_ISL_5990373 |
| hCoV-19/USA/IN-PU-lh000080651/2021 | EPI_ISL_5990374 |
| hCoV-19/USA/IN-PU-lh000098876/2021 | EPI_ISL_5990375 |
| hCoV-19/USA/IN-PU-lh000080906/2021 | EPI_ISL_5990376 |

| | |
|---|---|
| hCoV-19/USA/IN-PU-lh000094048/2021 | EPI_ISL_5990377 |
| hCoV-19/USA/IN-PU-lh000094260/2021 | EPI_ISL_5990378 |
| hCoV-19/USA/IN-PU-lh000094337/2021 | EPI_ISL_5990379 |
| hCoV-19/USA/IN-PU-lh000094066/2021 | EPI_ISL_5990380 |
| hCoV-19/USA/IN-PU-lh000132596/2021 | EPI_ISL_5990381 |
| hCoV-19/USA/IN-PU-lh000121584/2021 | EPI_ISL_5990382 |
| hCoV-19/USA/IN-PU-lh000121717/2021 | EPI_ISL_5990383 |
| hCoV-19/USA/IN-PU-lh000121568/2021 | EPI_ISL_5990384 |
| hCoV-19/USA/IN-PU-lh000121510/2021 | EPI_ISL_5990385 |
| hCoV-19/USA/IN-PU-lh000132664/2021 | EPI_ISL_5990386 |
| hCoV-19/USA/IN-PU-lh000132651/2021 | EPI_ISL_5990387 |
| hCoV-19/USA/IN-PU-lh000132996/2021 | EPI_ISL_5990388 |
| hCoV-19/USA/IN-PU-lh000132673/2021 | EPI_ISL_5990389 |
| hCoV-19/USA/IN-PU-lh000132975/2021 | EPI_ISL_5990390 |
| hCoV-19/USA/IN-PU-lh000094278/2021 | EPI_ISL_5990391 |
| hCoV-19/USA/IN-PU-lh000100042/2021 | EPI_ISL_5990392 |
| hCoV-19/USA/IN-PU-lh000094168/2021 | EPI_ISL_5990393 |
| hCoV-19/USA/IN-PU-lh000110618/2021 | EPI_ISL_5990394 |
| hCoV-19/USA/IN-PU-lh000082626/2021 | EPI_ISL_5990395 |
| hCoV-19/USA/IN-PU-lh000110798/2021 | EPI_ISL_5990396 |
| hCoV-19/USA/IN-PU-lh000082744/2021 | EPI_ISL_5990397 |
| hCoV-19/USA/IN-PU-lh000092563/2021 | EPI_ISL_5990398 |
| hCoV-19/USA/IN-PU-lh000080963/2021 | EPI_ISL_5990399 |
| hCoV-19/USA/IN-PU-lh000110644/2021 | EPI_ISL_5990401 |
| hCoV-19/USA/IN-PU-lh000124822/2021 | EPI_ISL_5990402 |
| hCoV-19/USA/IN-PU-lh000109243/2021 | EPI_ISL_5990403 |
| hCoV-19/USA/IN-PU-lh000082247/2021 | EPI_ISL_5990404 |

| | |
|---|---|
| hCoV-19/USA/IN-PU-lh000115021/2021 | EPI_ISL_5990405 |
| hCoV-19/USA/IN-PU-lh000145111/2021 | EPI_ISL_5990406 |
| hCoV-19/USA/IN-PU-lh000124985/2021 | EPI_ISL_5990407 |
| hCoV-19/USA/IN-PU-lh000110724/2021 | EPI_ISL_5990408 |
| hCoV-19/USA/IN-PU-lh000086762/2021 | EPI_ISL_5990409 |
| hCoV-19/USA/IN-PU-lh000080575/2021 | EPI_ISL_5990410 |
| hCoV-19/USA/IN-PU-lh000124798/2021 | EPI_ISL_5990411 |
| hCoV-19/USA/IN-PU-lh000124545/2021 | EPI_ISL_5990412 |
| hCoV-19/USA/IN-PU-lh000092223/2021 | EPI_ISL_5990413 |
| hCoV-19/USA/IN-PU-lh000116172/2021 | EPI_ISL_5990414 |
| hCoV-19/USA/IN-PU-lh000094127/2021 | EPI_ISL_5990415 |
| hCoV-19/USA/IN-PU-lh000082059/2021 | EPI_ISL_5990417 |
| hCoV-19/USA/IN-PU-lh000116157/2021 | EPI_ISL_5990418 |
| hCoV-19/USA/IN-PU-lh000145061/2021 | EPI_ISL_5990419 |
| hCoV-19/USA/IN-PU-lh000145070/2021 | EPI_ISL_5990420 |
| hCoV-19/USA/IN-PU-lh000145187/2021 | EPI_ISL_5990422 |
| hCoV-19/USA/IN-PU-lh000145177/2021 | EPI_ISL_5990424 |
| hCoV-19/USA/IN-PU-lh000145329/2021 | EPI_ISL_5990425 |
| hCoV-19/USA/IN-PU-lh000082161/2021 | EPI_ISL_5990426 |
| hCoV-19/USA/IN-PU-lh000082130/2021 | EPI_ISL_5990427 |
| hCoV-19/USA/IN-PU-lh000082070/2021 | EPI_ISL_5990428 |
| hCoV-19/USA/IN-PU-lh000080524/2021 | EPI_ISL_5990429 |
| hCoV-19/USA/IN-PU-lh000128781/2021 | EPI_ISL_5990431 |
| hCoV-19/USA/IN-PU-lh000100888/2021 | EPI_ISL_5990432 |
| hCoV-19/USA/IN-PU-lh000145805/2021 | EPI_ISL_5990433 |
| hCoV-19/USA/IN-PU-lh000082125/2021 | EPI_ISL_9272272 |
| hCoV-19/USA/IN-PU-lh000139869/2021 | EPI_ISL_9272260 |

| | |
|---|---|
| hCoV-19/USA/IN-PU-lh000105755/2021 | EPI_ISL_9272263 |
| hCoV-19/USA/IN-PU-lh000132705/2021 | EPI_ISL_9272259 |
| hCoV-19/USA/IN-PU-lh000133586/2021 | EPI_ISL_9272261 |
| hCoV-19/USA/IN-PU-lh000109361/2021 | EPI_ISL_9272275 |
| hCoV-19/USA/IN-PU-lh000116556/2021 | EPI_ISL_9272258 |
| hCoV-19/USA/IN-PU-lh000082300/2021 | EPI_ISL_9272276 |
| hCoV-19/USA/IN-PU-lh000086889/2021 | EPI_ISL_9272274 |
| hCoV-19/USA/IN-PU-lh000094379/2021 | EPI_ISL_9272270 |
| hCoV-19/USA/IN-PU-lh000086772/2021 | EPI_ISL_9272267 |
| hCoV-19/USA/IN-PU-LH000175526/2021 | EPI_ISL_9272330 |
| hCoV-19/USA/IN-PU-LH000091753/2021 | EPI_ISL_9272325 |
| hCoV-19/USA/IN-PU-LH000153280/2021 | EPI_ISL_9272324 |
| hCoV-19/USA/IN-PU-LH000157851/2021 | EPI_ISL_9272323 |
| hCoV-19/USA/IN-PU-LH000174039/2021 | EPI_ISL_9272329 |
| hCoV-19/USA/IN-PU-LH000111857/2021 | EPI_ISL_9272308 |
| hCoV-19/USA/IN-PU-LH000185968/2021 | EPI_ISL_9272301 |
| hCoV-19/USA/IN-PU-LH000183970/2021 | EPI_ISL_9272300 |
| hCoV-19/USA/IN-PU-LH000115719/2021 | EPI_ISL_9272289 |
| hCoV-19/USA/IN-PU-LH000172505/2021 | EPI_ISL_9272288 |
| hCoV-19/USA/IN-PU-LH000172528/2021 | EPI_ISL_9272296 |
| hCoV-19/USA/IN-PU-LH000043501/2021 | EPI_ISL_9272279 |
| hCoV-19/USA/IN-PU-LH000191709/2021 | EPI_ISL_9272292 |
| hCoV-19/USA/IN-PU-LH000176701/2021 | EPI_ISL_9272290 |
| hCoV-19/USA/IN-PU-LH000177062/2021 | EPI_ISL_9272278 |
| hCoV-19/USA/IN-PU-LH000177980/2021 | EPI_ISL_9272280 |
| hCoV-19/USA/IN-PU-LH000171893/2021 | EPI_ISL_9272298 |
| hCoV-19/USA/IN-PU-LH000184528/2021 | EPI_ISL_9272282 |

| | |
|---|---|
| hCoV-19/USA/IN-PU-LH000084431/2021 | EPI_ISL_9272283 |
| hCoV-19/USA/IN-PU-LH000175315/2021 | EPI_ISL_9272286 |
| hCoV-19/USA/IN-PU-LH000171072/2021 | EPI_ISL_9272287 |
| hCoV-19/USA/IN-PU-lh000133297/2021 | EPI_ISL_9272262 |
| hCoV-19/USA/IN-PU-LH000144729/2021 | EPI_ISL_9272335 |
| hCoV-19/USA/IN-PU-LH000189903/2021 | EPI_ISL_9272333 |
| hCoV-19/USA/IN-PU-LH000189345/2021 | EPI_ISL_9272332 |
| hCoV-19/USA/IN-PU-LH000111954/2021 | EPI_ISL_9272334 |
| hCoV-19/USA/IN-PU-LH000115829/2021 | EPI_ISL_9272281 |
| hCoV-19/USA/IN-PU-LH000110949/2021 | EPI_ISL_9272340 |
| hCoV-19/USA/IN-PU-LH000144906/2021 | EPI_ISL_9272337 |
| hCoV-19/USA/IN-PU-LH000166661/2021 | EPI_ISL_9272327 |
| hCoV-19/USA/IN-PU-LH000188890/2021 | EPI_ISL_9272328 |
| hCoV-19/USA/IN-PU-LH000155753/2021 | EPI_ISL_9272326 |
| hCoV-19/USA/IN-PU-LH000174492/2021 | EPI_ISL_9272331 |
| hCoV-19/USA/IN-PU-LH000101959/2021 | EPI_ISL_9272338 |
| hCoV-19/USA/IN-PU-LH000130564/2021 | EPI_ISL_9272339 |
| hCoV-19/USA/IN-PU-LH000143185/2021 | EPI_ISL_9272336 |
| hCoV-19/USA/IN-PU-LH000161550/2021 | EPI_ISL_9272321 |
| hCoV-19/USA/IN-PU-LH000116050/2021 | EPI_ISL_9272341 |
| hCoV-19/USA/IN-PU-LH000128241/2021 | EPI_ISL_9272317 |
| hCoV-19/USA/IN-PU-LH000111767/2021 | EPI_ISL_9272312 |
| hCoV-19/USA/IN-PU-LH000128194/2021 | EPI_ISL_9272315 |
| hCoV-19/USA/IN-PU-LH000111937/2021 | EPI_ISL_9272310 |
| hCoV-19/USA/IN-PU-LH000128469/2021 | EPI_ISL_9272311 |
| hCoV-19/USA/IN-PU-LH000155814/2021 | EPI_ISL_9272322 |
| hCoV-19/USA/IN-PU-LH000161626/2021 | EPI_ISL_9272320 |

| | |
|---|---|
| hCoV-19/USA/IN-PU-LH000160817/2021 | EPI_ISL_9272309 |
| hCoV-19/USA/IN-PU-LH000165147/2021 | EPI_ISL_9272318 |
| hCoV-19/USA/IN-PU-LH000147291/2021 | EPI_ISL_9272314 |
| hCoV-19/USA/IN-PU-LH000161578/2021 | EPI_ISL_9272319 |
| hCoV-19/USA/IN-PU-LH000128236/2021 | EPI_ISL_9272313 |
| hCoV-19/USA/IN-PU-LH000105661/2021 | EPI_ISL_9272306 |
| hCoV-19/USA/IN-PU-LH000162079/2021 | EPI_ISL_9272316 |
| hCoV-19/USA/IN-PU-LH000180687/2021 | EPI_ISL_9272294 |
| hCoV-19/USA/IN-PU-LH000180757/2021 | EPI_ISL_9272297 |
| hCoV-19/USA/IN-PU-LH000185947/2021 | EPI_ISL_9272302 |
| hCoV-19/USA/IN-PU-LH000179713/2021 | EPI_ISL_9272293 |
| hCoV-19/USA/IN-PU-LH000101852/2021 | EPI_ISL_9272307 |
| hCoV-19/USA/IN-PU-LH000185918/2021 | EPI_ISL_9272305 |
| hCoV-19/USA/IN-PU-LH000168774/2021 | EPI_ISL_9272303 |
| hCoV-19/USA/IN-PU-LH000171923/2021 | EPI_ISL_9272299 |
| hCoV-19/USA/IN-PU-LH000190568/2021 | EPI_ISL_9272295 |
| hCoV-19/USA/IN-PU-LH000082742/2021 | EPI_ISL_9272304 |
| hCoV-19/USA/IN-PU-LH000184527/2021 | EPI_ISL_9272284 |
| hCoV-19/USA/IN-PU-LH000191089/2021 | EPI_ISL_9272291 |
| hCoV-19/USA/IN-PU-LH000171946/2021 | EPI_ISL_9272285 |
| hCoV-19/USA/IN-PU-lh000145186/2021 | EPI_ISL_9272277 |
| hCoV-19/USA/IN-PU-lh000109039/2021 | EPI_ISL_9272269 |
| hCoV-19/USA/IN-PU-lh000124573/2021 | EPI_ISL_9272271 |
| hCoV-19/USA/IN-PU-lh000080653/2021 | EPI_ISL_9272268 |
| hCoV-19/USA/IN-PU-lh000110722/2021 | EPI_ISL_9272266 |
| hCoV-19/USA/IN-PU-lh000101716/2021 | EPI_ISL_9272265 |
| hCoV-19/USA/IN-PU-lh000101714/2021 | EPI_ISL_9272264 |

| | |
|---|---|
| hCoV-19/USA/IN-PU-lh000094259/2021 | EPI_ISL_9272257 |
| hCoV-19/USA/IN-PU-lh000145413/2021 | EPI_ISL_9272273 |
| hCoV-19/USA/IN-PU-LH000181022/2021 | EPI_ISL_10031346 |
| hCoV-19/USA/IN-PU-LH000178558/2021 | EPI_ISL_10031347 |
| hCoV-19/USA/IN-PU-lh000080582/2021 | EPI_ISL_10031676 |
| hCoV-19/USA/IN-PU-LH000101699/2021 | EPI_ISL_10031677 |
| hCoV-19/USA/IN-PU-LH000122042/2021 | EPI_ISL_10031678 |

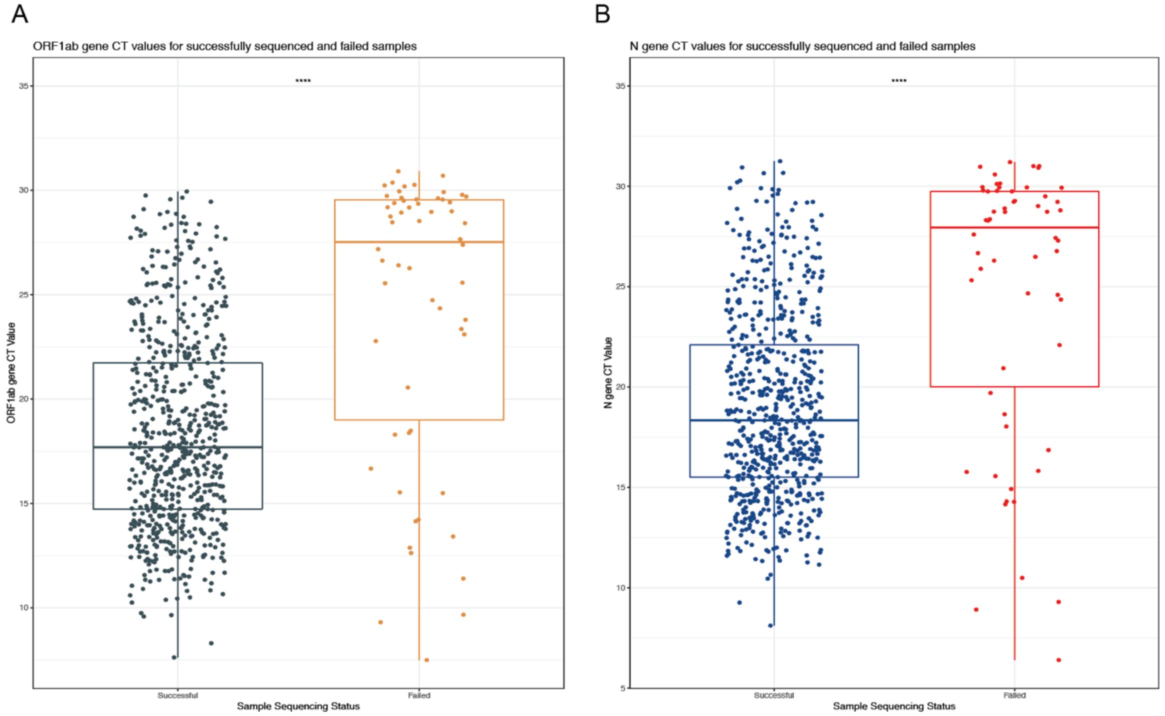


**Supplementary Figure 1.** Comparison of two SARS-CoV-2 genes targets of TaqPath RT-PCR for failed and successfully sequenced samples *(A)* Comparison of ORF1ab gene CT values among all samples that generated high or low-quality genomes. The first, second, and third quartiles are within the box, with the median line bolded. Samples are displayed as points with outliers included. P-values were generated using Wilcoxon rank-sum test and asterisks indicate P-value <0.05 (p<0.0001). *(B)* Comparison of N gene CT values among all samples that generated high or low-quality genomes. (S gene CT values are not included as a third comparison from RT-PCR target because SGTF presence or value of 0 would greatly affect comparisons).

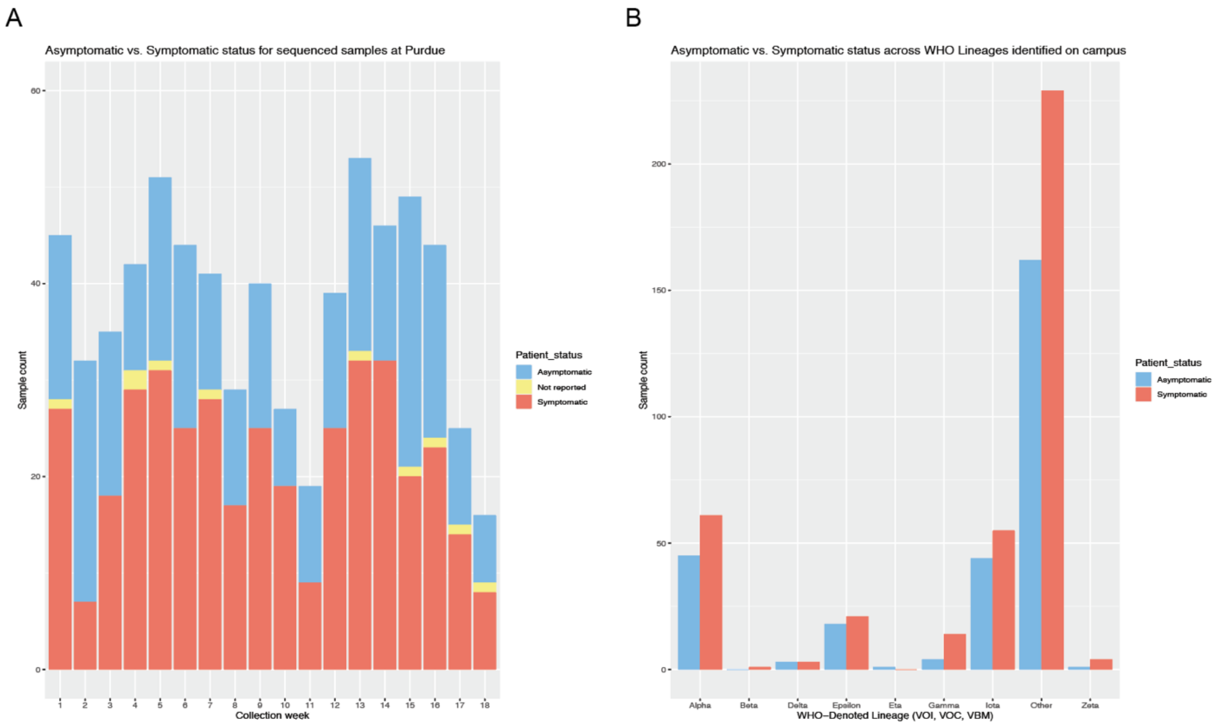


**Supplementary Figure 2.** Comparison of asymptomatic vs. symptomatic cases. *(A)* Barplot illustrating the distribution of asymptomatic vs. symptomatic patient status among sequenced cases across the 18-week study period. The x-axis displays collection weeks from January 3, 2021 to May 8, 2021, and the y-axis is the total sample count. *(B)* Barplot illustrating the distribution of asymptomatic vs. symptomatic patient status among sequenced cases by the WHO-identified lineages.

A

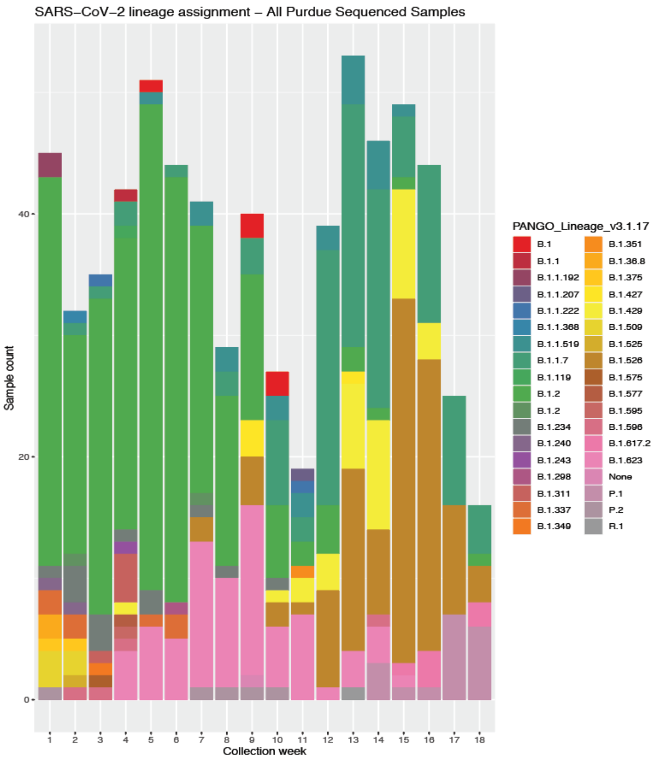


**Supplementary Figure 3.** Distribution of all SARS-CoV-2 variants on Purdue Campus. Barplot displays all identified SARS-CoV-2 variants from whole-genome sequenced samples each week from first week of January through the first week of May 2021. Each color represents a distinct variant as obtained from Pangolin version 3.1.17.
